## Supplementary Methods & Figures for "Estimating *R_e_* and overdispersion in secondary cases from the size of identical sequence clusters of SARS-CoV-2"

**11 Bibliography** **60**

### 1 Data

We used sequence data collected during the SARS-CoV-2 epidemic in Switzerland, Denmark and Germany to estimate the effective reproduction number $R_e$ and the dispersion parameter $k$ on a monthly basis during 2021. We downloaded all available sequences of the respective country and period from GISAID [1] and grouped them into clusters of identical sequences. A cluster is assigned to a month if there is a sequence in the cluster that has been sampled in the month. The same cluster can therefore be assigned to several months. This is why the clusters assigned to a month contain strictly more sequences than sequences were sampled during the respective month. Furthermore, we retrieved daily numbers of newly confirmed cases from public sources of Switzerland [2], Denmark [3] and Germany [4] and summarized them to monthly values. The number of confirmed cases, the number of sampled sequences, the number of assigned identical sequence clusters, the number of sequences contained in the assigned identical sequence clusters, the sequencing coverage and the size of the largest assigned identical sequence cluster by country and month are presented in the following tables.

#### 1.1 Switzerland

| Month | Nocc | Nos | Nosic | Noc | Nos/Nocc | Solc |
|---|---:|---:|---:|---:|---:|---:|
| January | 68,867 | 7,590 | 9,915 | 5,665 | 0.1102 | 114 |
| February | 32,110 | 4,213 | 6,367 | 2,712 | 0.1312 | 92 |
| March | 46,145 | 5,377 | 7,721 | 3,417 | 0.1165 | 92 |
| April | 58,135 | 7,485 | 10,226 | 4,957 | 0.1288 | 65 |
| May | 33,434 | 5,692 | 7,748 | 3,663 | 0.1702 | 65 |
| June | 7,381 | 1,586 | 3,335 | 1,178 | 0.2149 | 363 |
| July | 16,117 | 5,450 | 7,335 | 3,050 | 0.3382 | 363 |
| August | 64,669 | 12,456 | 15,772 | 8,389 | 0.1926 | 363 |
| September | 57,672 | 9,037 | 13,688 | 6,437 | 0.1567 | 363 |
| October | 35,422 | 9,030 | 13,967 | 5,848 | 0.2549 | 363 |
| November | 152,427 | 14,817 | 19,134 | 9,437 | 0.0972 | 204 |
| December | 324,389 | 10,700 | 16,277 | 7,989 | 0.0330 | 488 |
| Total | 896,768 | 93,433 | 131,485 | 62,742 | 0.1042 | 488 |

**Table S1. Overview of data from Switzerland.**
Nocc: Number of confirmed cases Nos: Number of sampled sequences Nosic: Number of sequences in clusters Noc: Number of clusters Solc: Size of largest clusters.

| | Cluster size | | | | | | | | | |
|---|---|---|---|---|---|---|---|---|---|---|
| **Month** | **1** | **2** | **3** | **4** | **5** | **6-10** | **11-50** | **51-100** | **101-488** | **All** |
| January | 4,482 | 574 | 223 | 104 | 63 | 125 | 90 | 3 | 1 | 5,665 |
| February | 1,872 | 345 | 146 | 82 | 53 | 116 | 96 | 2 | 0 | 2,712 |
| March | 2,421 | 428 | 160 | 98 | 61 | 138 | 108 | 3 | 0 | 3,417 |
| April | 3,537 | 639 | 262 | 140 | 87 | 168 | 121 | 3 | 0 | 4,957 |
| May | 2,602 | 451 | 192 | 107 | 67 | 146 | 96 | 2 | 0 | 3,663 |
| June | 833 | 144 | 63 | 34 | 21 | 48 | 29 | 4 | 2 | 1,178 |
| July | 2,231 | 405 | 138 | 77 | 35 | 81 | 70 | 10 | 3 | 3,050 |
| August | 6,493 | 995 | 334 | 179 | 95 | 166 | 110 | 13 | 4 | 8,389 |
| September | 4,755 | 819 | 318 | 155 | 93 | 170 | 110 | 13 | 4 | 6,437 |
| October | 4,179 | 707 | 309 | 146 | 93 | 242 | 157 | 11 | 4 | 5,848 |
| November | 7,044 | 1,083 | 429 | 233 | 140 | 302 | 193 | 11 | 2 | 9,437 |
| December | 6,274 | 801 | 301 | 156 | 97 | 190 | 153 | 12 | 5 | 7,989 |
| Total | 46,723 | 7,391 | 2,875 | 1,511 | 905 | 1,892 | 1,333 | 87 | 25 | 62,742 |

**Table S2A. Identical sequence clusters from Switzerland.**

| | Cluster size | | | | | | | | | |
|---|---|---|---|---|---|---|---|---|---|---|
| **Month** | **1** | **2** | **3** | **4** | **5** | **6-10** | **11-50** | **51-100** | **101-488** | **All** |
| January | 0.7912 | 0.1013 | 0.0394 | 0.0184 | 0.0111 | 0.0221 | 0.0159 | 0.0005 | 0.0002 | 1 |
| February | 0.6903 | 0.1272 | 0.0538 | 0.0302 | 0.0195 | 0.0428 | 0.0354 | 0.0007 | 0.0000 | 1 |
| March | 0.7085 | 0.1253 | 0.0468 | 0.0287 | 0.0179 | 0.0404 | 0.0316 | 0.0009 | 0.0000 | 1 |
| April | 0.7135 | 0.1289 | 0.0529 | 0.0282 | 0.0176 | 0.0339 | 0.0244 | 0.0006 | 0.0000 | 1 |
| May | 0.7103 | 0.1231 | 0.0524 | 0.0292 | 0.0183 | 0.0399 | 0.0262 | 0.0005 | 0.0000 | 1 |
| June | 0.7071 | 0.1222 | 0.0535 | 0.0289 | 0.0178 | 0.0407 | 0.0246 | 0.0034 | 0.0017 | 1 |
| July | 0.7315 | 0.1328 | 0.0452 | 0.0252 | 0.0115 | 0.0266 | 0.0230 | 0.0033 | 0.0010 | 1 |
| August | 0.7740 | 0.1186 | 0.0398 | 0.0213 | 0.0113 | 0.0198 | 0.0131 | 0.0015 | 0.0005 | 1 |
| September | 0.7387 | 0.1272 | 0.0494 | 0.0241 | 0.0144 | 0.0264 | 0.0171 | 0.0020 | 0.0006 | 1 |
| October | 0.7146 | 0.1209 | 0.0528 | 0.0250 | 0.0159 | 0.0414 | 0.0268 | 0.0019 | 0.0007 | 1 |
| November | 0.7464 | 0.1148 | 0.0455 | 0.0247 | 0.0148 | 0.0320 | 0.0205 | 0.0012 | 0.0002 | 1 |
| December | 0.7853 | 0.1003 | 0.0377 | 0.0195 | 0.0121 | 0.0238 | 0.0192 | 0.0015 | 0.0006 | 1 |
| Total | 0.7447 | 0.1178 | 0.0458 | 0.0241 | 0.0144 | 0.0302 | 0.0212 | 0.0014 | 0.0004 | 1 |

**Table S2B. Identical sequence clusters from Switzerland (proportion of clusters).**

#### 1.2 Denmark

| Month | Nocc | Nos | Nosic | Noc | Nos/Nocc | Solc |
|---|---|---|---|---|---|---|
| January | 32,069 | 14,784 | 29,882 | 5,273 | 0.4610 | 417 |
| February | 12,856 | 9,265 | 25,068 | 2,787 | 0.7207 | 926 |
| March | 19,131 | 16,159 | 27,755 | 4,196 | 0.8447 | 926 |
| April | 20,550 | 14,715 | 34,189 | 4,485 | 0.7161 | 926 |
| May | 29,713 | 23,142 | 35,922 | 6,077 | 0.7789 | 926 |
| June | 11,614 | 5,065 | 19,175 | 1,881 | 0.4361 | 760 |
| July | 23,788 | 19,769 | 28,362 | 7,459 | 0.8310 | 760 |
| August | 29,032 | 23,859 | 35,669 | 10,625 | 0.8218 | 518 |
| September | 12,528 | 10,767 | 23,179 | 5,102 | 0.8594 | 1,278 |
| October | 30,881 | 24,213 | 46,539 | 8,160 | 0.7841 | 1,278 |
| November | 106,733 | 49,645 | 77,490 | 18,872 | 0.4651 | 1,278 |
| December | 324,105 | 37,854 | 72,492 | 16,908 | 0.1168 | 1,278 |
| Total | 653,000 | 249,237 | 455,722 | 91,825 | 0.3817 | 1,278 |

**Table S3. Overview of data from Denmark.**
Nocc: Number of confirmed cases Nos: Number of sampled sequences Nosic: Number of sequences in clusters Noc: Number of clusters Solc: Size of largest clusters.

| | Cluster size | | | | | | | | | |
|---|---|---|---|---|---|---|---|---|---|---|
| **Month** | **1** | **2** | **3** | **4** | **5** | **6-10** | **11-50** | **51-100** | **101-1278** | **All** |
| January | 3,238 | 663 | 312 | 174 | 131 | 299 | 359 | 48 | 49 | 5,273 |
| February | 1,595 | 371 | 157 | 94 | 81 | 168 | 231 | 37 | 53 | 2,787 |
| March | 2,499 | 555 | 305 | 138 | 104 | 242 | 264 | 41 | 48 | 4,196 |
| April | 2,629 | 633 | 275 | 152 | 116 | 254 | 304 | 65 | 57 | 4,485 |
| May | 3,902 | 789 | 319 | 186 | 126 | 296 | 342 | 66 | 51 | 6,077 |
| June | 1,148 | 237 | 115 | 60 | 36 | 90 | 121 | 31 | 43 | 1,881 |
| July | 5,140 | 965 | 388 | 233 | 122 | 309 | 229 | 39 | 34 | 7,459 |
| August | 6,945 | 1,534 | 624 | 332 | 220 | 513 | 379 | 47 | 31 | 10,625 |
| September | 3,256 | 643 | 298 | 194 | 121 | 259 | 270 | 36 | 25 | 5,102 |
| October | 5,126 | 1,087 | 482 | 256 | 161 | 428 | 447 | 101 | 72 | 8,160 |
| November | 12,843 | 2,230 | 935 | 504 | 338 | 928 | 868 | 143 | 83 | 18,872 |
| December | 11,490 | 2,021 | 874 | 480 | 289 | 800 | 744 | 115 | 95 | 16,908 |
| Total | 59,811 | 11,728 | 5,084 | 2,803 | 1,845 | 4,586 | 4,558 | 769 | 641 | 91,825 |

**Table S4A. Identical sequence clusters from Denmark.**

| | Cluster size | | | | | | | | | |
|---|---|---|---|---|---|---|---|---|---|---|
| **Month** | **1** | **2** | **3** | **4** | **5** | **6-10** | **11-50** | **51-100** | **101-1278** | **All** |
| January | 0.6141 | 0.1257 | 0.0592 | 0.0330 | 0.0248 | 0.0567 | 0.0681 | 0.0091 | 0.0093 | 1 |
| February | 0.5723 | 0.1331 | 0.0563 | 0.0337 | 0.0291 | 0.0603 | 0.0829 | 0.0133 | 0.0190 | 1 |
| March | 0.5956 | 0.1323 | 0.0727 | 0.0329 | 0.0248 | 0.0577 | 0.0629 | 0.0098 | 0.0114 | 1 |
| April | 0.5862 | 0.1411 | 0.0613 | 0.0339 | 0.0259 | 0.0566 | 0.0678 | 0.0145 | 0.0127 | 1 |
| May | 0.6421 | 0.1298 | 0.0525 | 0.0306 | 0.0207 | 0.0487 | 0.0563 | 0.0109 | 0.0084 | 1 |
| June | 0.6103 | 0.1260 | 0.0611 | 0.0319 | 0.0191 | 0.0478 | 0.0643 | 0.0165 | 0.0229 | 1 |
| July | 0.6891 | 0.1294 | 0.0520 | 0.0312 | 0.0164 | 0.0414 | 0.0307 | 0.0052 | 0.0046 | 1 |
| August | 0.6536 | 0.1444 | 0.0587 | 0.0312 | 0.0207 | 0.0483 | 0.0357 | 0.0044 | 0.0029 | 1 |
| September | 0.6382 | 0.1260 | 0.0584 | 0.0380 | 0.0237 | 0.0508 | 0.0529 | 0.0071 | 0.0049 | 1 |
| October | 0.6282 | 0.1332 | 0.0591 | 0.0314 | 0.0197 | 0.0525 | 0.0548 | 0.0124 | 0.0088 | 1 |
| November | 0.6805 | 0.1182 | 0.0495 | 0.0267 | 0.0179 | 0.0492 | 0.0460 | 0.0076 | 0.0044 | 1 |
| December | 0.6796 | 0.1195 | 0.0517 | 0.0284 | 0.0171 | 0.0473 | 0.0440 | 0.0068 | 0.0056 | 1 |
| Total | 0.6514 | 0.1277 | 0.0554 | 0.0305 | 0.0201 | 0.0499 | 0.0496 | 0.0084 | 0.0070 | 1 |

**Table S4B. Identical sequence clusters from Denmark (proportion of clusters).**

##### 1.3 Germany

| Month | Nocc | Nos | Nosic | Noc | Nos/Nocc | Solc |
|---|---:|---:|---:|---:|---:|---:|
| January | 482,531 | 6,860 | 10,872 | 5,024 | 0.0142 | 176 |
| February | 224,866 | 18,287 | 28,013 | 11,870 | 0.0813 | 183 |
| March | 373,570 | 34,372 | 49,996 | 20,202 | 0.0920 | 226 |
| April | 567,343 | 41,564 | 59,711 | 25,764 | 0.0733 | 226 |
| May | 294,982 | 26,786 | 42,130 | 18,234 | 0.0908 | 226 |
| June | 48,307 | 8,155 | 13,741 | 5,668 | 0.1688 | 283 |
| July | 43,089 | 9,001 | 12,960 | 5,687 | 0.2089 | 283 |
| August | 178,357 | 29,427 | 39,608 | 20,224 | 0.1650 | 283 |
| September | 284,190 | 40,701 | 54,270 | 27,844 | 0.1432 | 283 |
| October | 377,348 | 31,376 | 46,176 | 21,753 | 0.0831 | 283 |
| November | 1,255,833 | 47,177 | 62,969 | 34,850 | 0.0376 | 283 |
| December | 1,299,987 | 51,047 | 71,458 | 38,833 | 0.0393 | 800 |
| Total | 5,430,403 | 344,753 | 491,904 | 235,953 | 0.0635 | 800 |

**Table S5. Overview of data from Germany.**
Nocc: Number of confirmed cases Nos: Number of sampled sequences Nosic: Number of sequences in clusters Noc: Number of clusters Solc: Size of largest clusters.

| | Cluster size | | | | | | | | | |
|---|---|---|---|---|---|---|---|---|---|---|
| **Month** | **1** | **2** | **3** | **4** | **5** | **6-10** | **11-50** | **51-100** | **101-800** | **All** |
| January | 3,779 | 554 | 215 | 129 | 88 | 142 | 102 | 12 | 3 | 5,024 |
| February | 8,568 | 1,418 | 579 | 312 | 223 | 405 | 331 | 27 | 7 | 11,870 |
| March | 14,022 | 2,557 | 1,077 | 604 | 400 | 879 | 610 | 42 | 11 | 20,202 |
| April | 17,982 | 3,301 | 1,420 | 795 | 503 | 1,050 | 665 | 37 | 11 | 25,764 |
| May | 12,747 | 2,405 | 1,015 | 519 | 352 | 698 | 464 | 24 | 10 | 18,234 |
| June | 3,925 | 784 | 302 | 164 | 120 | 225 | 128 | 16 | 4 | 5,668 |
| July | 4,060 | 772 | 274 | 144 | 94 | 196 | 134 | 8 | 5 | 5,687 |
| August | 15,006 | 2,527 | 987 | 464 | 315 | 551 | 348 | 17 | 9 | 20,224 |
| September | 20,418 | 3,617 | 1,353 | 716 | 459 | 781 | 468 | 23 | 9 | 27,844 |
| October | 15,817 | 2,667 | 1,029 | 571 | 391 | 770 | 475 | 25 | 8 | 21,753 |
| November | 26,384 | 4,405 | 1,465 | 783 | 469 | 846 | 466 | 24 | 8 | 34,850 |
| December | 30,802 | 4,163 | 1,432 | 740 | 398 | 735 | 503 | 38 | 22 | 38,833 |
| Total | 173,510 | 29,170 | 11,148 | 5,941 | 3,812 | 7,278 | 4,694 | 293 | 107 | 235,953 |

**Table S6A. Identical sequence clusters from Germany.**

| | Cluster size | | | | | | | | | |
|---|---|---|---|---|---|---|---|---|---|---|
| **Month** | **1** | **2** | **3** | **4** | **5** | **6-10** | **11-50** | **51-100** | **101-800** | **All** |
| January | 0.7522 | 0.1103 | 0.0428 | 0.0257 | 0.0175 | 0.0283 | 0.0203 | 0.0024 | 0.0006 | 1 |
| February | 0.7218 | 0.1195 | 0.0488 | 0.0263 | 0.0188 | 0.0341 | 0.0279 | 0.0023 | 0.0006 | 1 |
| March | 0.6941 | 0.1266 | 0.0533 | 0.0299 | 0.0198 | 0.0435 | 0.0302 | 0.0021 | 0.0005 | 1 |
| April | 0.6980 | 0.1281 | 0.0551 | 0.0309 | 0.0195 | 0.0408 | 0.0258 | 0.0014 | 0.0004 | 1 |
| May | 0.6991 | 0.1319 | 0.0557 | 0.0285 | 0.0193 | 0.0383 | 0.0254 | 0.0013 | 0.0005 | 1 |
| June | 0.6925 | 0.1383 | 0.0533 | 0.0289 | 0.0212 | 0.0397 | 0.0226 | 0.0028 | 0.0007 | 1 |
| July | 0.7139 | 0.1357 | 0.0482 | 0.0253 | 0.0165 | 0.0345 | 0.0236 | 0.0014 | 0.0009 | 1 |
| August | 0.7420 | 0.1250 | 0.0488 | 0.0229 | 0.0156 | 0.0272 | 0.0172 | 0.0008 | 0.0004 | 1 |
| September | 0.7333 | 0.1299 | 0.0486 | 0.0257 | 0.0165 | 0.0280 | 0.0168 | 0.0008 | 0.0003 | 1 |
| October | 0.7271 | 0.1226 | 0.0473 | 0.0262 | 0.0180 | 0.0354 | 0.0218 | 0.0011 | 0.0004 | 1 |
| November | 0.7571 | 0.1264 | 0.0420 | 0.0225 | 0.0135 | 0.0243 | 0.0134 | 0.0007 | 0.0002 | 1 |
| December | 0.7932 | 0.1072 | 0.0369 | 0.0191 | 0.0102 | 0.0189 | 0.0130 | 0.0010 | 0.0006 | 1 |
| Total | 0.7354 | 0.1236 | 0.0472 | 0.0252 | 0.0162 | 0.0308 | 0.0199 | 0.0012 | 0.0005 | 1 |

**Table S6B. Identical sequence clusters from Germany (proportion of clusters).**

### 2 Probability Theory

In this chapter, we present a detailed mathematical derivation of a statement upon which our model of the size distribution of identical sequence clusters is built: If the number of secondary cases created by an infected individual follows a negative binomial distribution with mean˜$R_e$ and dispersion parameter˜$k$, then the number of secondary cases that belong to the same identical sequence cluster as their source case also follows a negative binomial distribution with mean $R_g = (1-\mu)R_e$, where $\mu$ is the probability of a mutation occurring in an infected individual, and dispersion parameter $k$.

First, we present this result in a mathematical context. Its application within our model of the size distribution of identical sequence clusters will be explained in the next chapter. We start by proofing two auxiliary results that we will use within the proof of our main statement.

**Lemma 1.** *For all $R, k \in \mathbb{R}_{>0}$, for all $\mu \in [0,1]$ and for all $j \in \mathbb{Z}_{\geq 0}$:*

$$\sum_{l=0}^{\infty} \frac{\Gamma(k+j+l)}{l!\Gamma(k+j)} \left(\frac{\mu R}{k+R}\right)^l = \left(\frac{k+R}{k+(1-\mu)R}\right)^{k+j}.$$

*Proof.* We define

$$f_j(x) = x^{-(k+j)}.$$

By Taylor's theorem:

$$\begin{aligned}
\left(\frac{k+R}{k+(1-\mu)R}\right)^{k+j} &= f_j\left(\frac{k+(1-\mu)R}{k+R}\right) \\
&= f_j(1) + \sum_{l=1}^{\infty} \frac{f_j^{(l)}(1)}{l!} \left(\frac{k+(1-\mu)R}{k+R} - 1\right)^l \\
&= 1 + \sum_{l=1}^{\infty} \frac{\prod_{m=0}^{l-1} -(k+j+m)}{l!} \left(-\frac{\mu R}{k+R}\right)^l \\
&= 1 + \sum_{l=1}^{\infty} \frac{\prod_{m=0}^{l-1} (k+j+m)}{l!} \left(\frac{\mu R}{k+R}\right)^l \\
&= \sum_{l=0}^{\infty} \frac{\Gamma(k+j+l)}{l!\Gamma(k+j)} \left(\frac{\mu R}{k+R}\right)^l
\end{aligned}$$

□

**Lemma 2.** *For all $R, k \in \mathbb{R}_{>0}$, for all $\mu \in [0,1]$ and for all $j \in \mathbb{Z}_{\geq 0}$:*

$$\sum_{i=j}^{\infty} \frac{\Gamma(k+i)}{(i-j)!\Gamma(k+j)} (\mu R)^{i-j} (k+(1-\mu)R)^{k+j} (k+R)^{-(k+i)} = 1$$

*Proof.* It holds:

$$\begin{aligned}
&\sum_{i=j}^{\infty} \frac{\Gamma(k+i)}{(i-j)!\Gamma(k+j)} (\mu R)^{i-j} (k+(1-\mu)R)^{k+j} (k+R)^{-(k+i)} \\
&= \left(\frac{k+(1-\mu)R}{k+R}\right)^{k} \sum_{i=j}^{\infty} \frac{\Gamma(k+i)}{(i-j)!\Gamma(k+j)} (\mu R)^{i-j} (k+(1-\mu)R)^{j} (k+R)^{-i} \\
&= \left(\frac{k+(1-\mu)R}{k+R}\right)^{k} \sum_{l=0}^{\infty} \frac{\Gamma(k+j+l)}{l!\Gamma(k+j)} (\mu R)^{l} (k+(1-\mu)R)^{j} (k+R)^{-(j+l)} \\
&= \left(\frac{k+(1-\mu)R}{k+R}\right)^{k} \sum_{l=0}^{\infty} \frac{\Gamma(k+j+l)}{l!\Gamma(k+j)} \left(\frac{k+(1-\mu)R}{k+R}\right)^{j} \left(\frac{\mu R}{k+R}\right)^{l} \\
&= \left(\frac{k+(1-\mu)R}{k+R}\right)^{k+j} \sum_{l=0}^{\infty} \frac{\Gamma(k+j+l)}{l!\Gamma(k+j)} \left(\frac{\mu R}{k+R}\right)^{l}
\end{aligned}$$

We conclude by applying Lemma 1. □

Next, we turn to the proof of the main statement of this chapter.

**Theorem 1.** *Let $R, k \in \mathbb{R}_{>0}$ and let $\mu \in [0,1]$. Furthermore, let $X$ be an integer-valued random variable following a negative binomial distribution with mean $R$ and dispersion parameter $k$,*

$$\text{i.e.} \quad \mathbb{P}\left(X=i\right)=\frac{\Gamma\left(k+i\right)}{i!\Gamma\left(k\right)}\left(\frac{k}{k+R}\right)^{k}\left(\frac{R}{k+R}\right)^{i} \quad \text{for all}\, i \in \mathbb{Z}_{\geq 0}.$$

*The discrete random variable $Y$ with*

$$\mathbb{P}\left(Y=j\right)=\sum_{i=j}^{\infty}\binom{i}{j}\left(1-\mu\right)^{j}\mu^{i-j}\,\mathbb{P}\left(X=i\right) \quad \text{for all}\, j \in \mathbb{Z}_{\geq 0}$$

*follows a negative binomial distribution with mean $\left(1-\mu\right)R$ and dispersion parameter $k$.*

*Proof.* For all $j \in \mathbb{Z}_{\geq 0}$ we get:

$$\begin{aligned}
\mathbb{P}\left(Y=j\right) &= \sum_{i=j}^{\infty}\binom{i}{j}\left(1-\mu\right)^{j}\mu^{i-j}\,\mathbb{P}\left(X=i\right)\\
&= \sum_{i=j}^{\infty}\binom{i}{j}\left(1-\mu\right)^{j}\mu^{i-j}\frac{\Gamma\left(k+i\right)}{i!\Gamma\left(k\right)}\left(\frac{k}{k+R}\right)^{k}\left(\frac{R}{k+R}\right)^{i}\\
&= \left(\frac{k}{k+\left(1-\mu\right)R}\right)^{k}\sum_{i=j}^{\infty}\binom{i}{j}\left(1-\mu\right)^{j}\mu^{i-j}\frac{\Gamma\left(k+i\right)}{i!\Gamma\left(k\right)}\left(\frac{k}{k+R}\right)^{k}\left(\frac{k+\left(1-\mu\right)R}{k}\right)^{k}\left(\frac{R}{k+R}\right)^{i}\\
&= \left(\frac{k}{k+\left(1-\mu\right)R}\right)^{k}\sum_{i=j}^{\infty}\binom{i}{j}\left(1-\mu\right)^{j}\mu^{i-j}\frac{\Gamma\left(k+i\right)}{i!\Gamma\left(k\right)}\left(\frac{k+\left(1-\mu\right)R}{k+R}\right)^{k}\left(\frac{R}{k+R}\right)^{i}\\
&= \frac{\Gamma\left(k+j\right)}{j!\Gamma\left(k\right)}\left(\frac{k}{k+\left(1-\mu\right)R}\right)^{k}\sum_{i=j}^{\infty}\left(1-\mu\right)^{j}\mu^{i-j}\frac{\Gamma\left(k+i\right)}{\left(i-j\right)!\Gamma\left(k+j\right)}\left(\frac{k+\left(1-\mu\right)R}{k+R}\right)^{k}\left(\frac{R}{k+R}\right)^{i}
\end{aligned}$$

Furthermore,

$$\begin{aligned}
&\sum_{i=j}^{\infty}\left(1-\mu\right)^{j}\mu^{i-j}\frac{\Gamma\left(k+i\right)}{\left(i-j\right)!\Gamma\left(k+j\right)}\left(\frac{k+\left(1-\mu\right)R}{k+R}\right)^{k}\left(\frac{R}{k+R}\right)^{i}\\
&= \left(\frac{\left(1-\mu\right)R}{k+\left(1-\mu\right)R}\right)^{j}\sum_{i=j}^{\infty}\mu^{i-j}\frac{\Gamma\left(k+i\right)}{\left(i-j\right)!\Gamma\left(k+j\right)}\left(\frac{k+\left(1-\mu\right)R}{k+R}\right)^{k}\left(\frac{k+\left(1-\mu\right)R}{R}\right)^{j}\left(\frac{R}{k+R}\right)^{i}\\
&= \left(\frac{\left(1-\mu\right)R}{k+\left(1-\mu\right)R}\right)^{j}\sum_{i=j}^{\infty}\frac{\Gamma\left(k+i\right)}{\left(i-j\right)!\Gamma\left(k+j\right)}\left(\mu R\right)^{i-j}\left(k+\left(1-\mu\right)R\right)^{k+j}\left(k+R\right)^{-\left(k+i\right)}
\end{aligned}$$

By Lemma 2, we get

$$\mathbb{P}\left(Y=j\right)=\frac{\Gamma\left(k+j\right)}{j!\Gamma\left(k\right)}\left(\frac{k}{k+\left(1-\mu\right)R}\right)^{k}\left(\frac{\left(1-\mu\right)R}{k+\left(1-\mu\right)R}\right)^{j}$$

□

### 3 Model of cluster size distribution

Both for inferring the effective reproduction number $R_e$ and the dispersion parameter $k$ from genomic sequence data and for simulating clusters of identical sequences we built upon a classical branching process setup that we applied to viral transmission. Starting with one source case, the cases of the subsequent generation are determined by a recurrence relation. Ordering and connecting the cases according to their generation and direct predecessor, we can represent the disease outbreak as a graph, more precisely as a rooted tree. This is why in this paragraph we often use the language of graph theory, denoting cases as nodes, or all cases sharing a common direct predecessor as offspring of a node.

We start with the same assumption that Blumberg and Lloyd-Smith [5] have made: For each node, the number of offspring $T$ follows a negative binomial distribution with mean $R_e$ and dispersion parameter $k$,

$$\mathbb{P}(T=i)=\frac{\Gamma(k+i)}{i!\,\Gamma(k)}\left(\frac{k}{k+R_e}\right)^k\left(\frac{R_e}{k+R_e}\right)^i \quad \text{for all } i\in\mathbb{Z}_{\geq 0}.$$

Having set up the scaffolding of our transmission model, we move to integrating the mutation process of the virus to separate different viral clades. At each node the occurrence of a mutation is modelled by a random variable following a Bernoulli distribution with parameter $\mu$. We assume that at each node a mutation occurs independently of all other nodes and prior to the respective node transmitting the virus to secondary cases or being detected. Therefore, if a mutation happens at a node, all direct offspring of that particular node receive the mutation. We use two different approaches to estimate $\mu$. On the one hand, we use an estimate of $\mu$ that is based on the estimation of the mean number of mutations per transmission of Park et al. [6]. Based on 87 transmission pairs, they obtained an estimate of $M_T = 0.33$ mutations per transmission on average with a 95% confidence interval of $0.22 - 0.48$. Plugging $M_T = 0.33$ into the formula

$$\mu = 1 - \mathrm{e}^{-M_T},$$

we get $\mu = 28.1\%$. On the other hand, we determined the mutation probability $\mu$ as

$$\mu = 1 - \mathrm{e}^{-MD/365.25},$$

where $M$ is the yearly mutation rate of SARS-CoV-2 and $D$ the mean generation interval. Based on the estimates of $M = 14$ mutations per year obtained by Neher et al. [7] and $D = 5.2$ days obtained by Ganyani et al. [8], we obtain $\mu = 18.1\%$.

In order to determine the distribution of the size of the identical sequence clusters, we need to determine the distribution of the number of offspring $V$ of a node that belong to the same identical sequence cluster as their parent. The number of offspring within the same identical sequence cluster of a node is the number of children of the node at which no mutation occurred, i.e. the genotype of the virus these children transmit to their offspring is identical to the genotype of the virus they received from their parent. Therefore,

$$\mathbb{P}(V=j)=\sum_{i=j}^{\infty}\binom{i}{j}(1-\mu)^j\,\mu^{i-j}\,\mathbb{P}(T=i) \quad \text{for all } j\in\mathbb{Z}_{\geq 0}.$$

$\mathbb{P}(V=j)$ is the probability that at $j$ children of a node no mutation occurs (and at all other children a mutation occurs), which means that these $j$ children belong to the same cluster as their parent. By Theorem 1, $V$ follows a negative binomial distribution with mean $R_g = (1-\mu)\,R_e$, the genomic reproduction number, and dispersion parameter $k$, i.e.,

$$\mathbb{P}(V=j)=\frac{\Gamma(k+j)}{j!\,\Gamma(k)}\left(\frac{k}{k+R_g}\right)^k\left(\frac{R_g}{k+R_g}\right)^j \quad \text{for all } j\in\mathbb{Z}_{\geq 0}.$$

Following Blumberg and Llyod-Smith [5], we determine the probability that an identical sequence cluster has size $j$ by

$$\mathbb{P}(X = j) = \frac{\Gamma(kj + j - 1)}{\Gamma(kj)\,\Gamma(j+1)} \frac{\left(\frac{R_g}{k}\right)^{j-1}}{\left(1 + \frac{R_g}{k}\right)^{kj+j-1}} \quad \text{for all } j \in \mathbb{Z}_{\geq 1}.$$

As did Blumberg and Llyod-Smith [9], we model the observation of each node by a random variable following a Bernoulli distribution with parameter $\tau$, the detection probability. The observation of each node is independent of the observation of all other nodes. The probability $\tau$ is determined as

$$\tau = \tau_{\text{test}}\ \tau_{\text{sequence}},$$

where $\tau_{\text{test}}$ is the probability that a case is confirmed by a test and $\tau_{\text{sequence}}$ is the probability that the genome of a confirmed case is sequenced. Consequently, the probability of observing $j$ cases contained in an identical sequence cluster of size $\geq j$ is given by

$$\mathbb{P}(Y = j) = \begin{cases} \sum_{i=1}^{\infty} (1-\tau)^i\ \mathbb{P}(X = i) & \text{if } j = 0, \\ \sum_{i=j}^{\infty} \binom{i}{j} \tau^j (1-\tau)^{i-j}\ \mathbb{P}(X = i) & \text{if } j \in \mathbb{Z}_{\geq 1}. \end{cases}$$

When implementing our model, we approximate the above infinite sums by limiting the number of summands to a finite threshold depending on $\tau$. For smaller values of $\tau$, the number of summands included is larger. As identical sequence clusters of which no case is observed do not appear in the data sets, the distribution of $Y$ does not appropriately model the size distribution of the identical sequence clusters contained in the data. To bridge this difference, we define the random variable $Z$, the number of observed cases in an identical sequence cluster of which at least one case has been observed. It holds:

$$\mathbb{P}(Z = j) = \frac{\mathbb{P}(Y = j)}{1 - \mathbb{P}(Y = 0)} \quad \text{for all } j \in \mathbb{Z}_{\geq 1}.$$

##### 3.1 Possible extensions of the model

If the genomic reproduction number $R_g$ is larger than 1, then the probability that an identical sequence cluster does not go extinct is strictly bigger than 0. This means that

$$\pi = \sum_{j=0}^{\infty} \mathbb{P}(Z = j) < 1.$$

Since we can not observe identical sequence clusters of arbitrary size, this requires additional adjustments to the model in order to fully cope with this situation. We have thought about possible approaches, but have not found a satisfactory solution. In the following we outline our ideas.

Nishiura et al. [10] have shown how to compute the probability of extinction of a branching process with a negative binomially distributed offspring distribution. Their arguments can be extended to compute˜$\pi$. Computing the size distribution of identical sequence clusters conditioned on them being finite is therefore possible but would require some additional computations. In our opinion, the bigger challenge lies elsewhere, namely in the classification of the potential of the clusters to grow to a large size.

Let us assume that $R_g > 1$ for a certain period. We have a number of clusters growing during that period, some already active before, some starting during this period. It is very likely that some of the clusters that have started during this period would continue to grow indefinitely if $R_g$ would stay above 1. In reality, this does not happen. $R_g$ will decrease below 1 at some point and all clusters will go extinct.

However, in our calculations to determine the cluster size distribution from the secondary case distribution, the effective reproduction number, the dispersion parameter, the mutation probability and the testing probability are assumed to remain constant. In order for the cluster size distribution with $R_g > 1$ derived from theory to be consistent with the observed cluster size distribution, one needs to know which clusters from the data would have grown to arbitrary size if $R_g$ stayed above 1. Similarly, to establish consistency between the cluster size distribution derived from theory conditioned on clusters going extinct with the data, one would need to remove those clusters from the data that would not go extinct if $R_g$ stayed above 1.

We have not managed to implement a method to separate clusters that would be likely to continue to grow if $R_g$ stayed above 1 from those that would go extinct. Therefore, we have left this question open for the time being. However, we are convinced that this would be an exciting starting point for further development of our model.

### 4 Prior distributions and fixed parameters

We set weakly informative prior distributions for the effective reproduction number $R_e$, the dispersion parameter $k$ and the testing probability $\tau_{\text{test}}$. Depending on the method the mutation process was included into the model, we either directly set a prior distribution for the mutation probability $\mu$ reflecting the uncertainty of the estimate obtained by Park et al. [6] or a tight prior distribution for the yearly mutation rate $M$, whose value was informed by work of Neher et al. [7]. We used the same prior distributions for all countries and months. For the prior distributions of $R_e$, $k$, $\mu$ and $M$ we opted for widely used probability distributions such as the gamma, beta or normal distribution. In addition, we incorporated expert knowledge about the proportion of new cases that are confirmed by a test into the prior distribution of $\tau_{\text{test}}$. We scaled a beta distribution, usually defined on the interval [0, 1] or ]0, 1[, to the interval [0.05, 1]. The reason for applying this transformation is to control the computational cost of our inference model. The lower $\tau_{test}$, the more summands are required to reliably approximate $\mathbb{P}(Y = j)$, see chapter Model of cluster size distribution for the definition of $Y$. Since we are convinced that at least 5% of all COVID-19 cases were confirmed by a test at any time in 2021 in Switzerland, Denmark and Germany, we have shifted the lower limit of the domain of the prior distribution of $\tau_{test}$ from 0 to 0.05. The probability density function of the resulting probability distribution, which we denote by scaled beta distribution, is given by the following formula:

$$f(x) = \frac{(x - 0.05)^{\alpha-1}(1-x)^{\beta-1}}{(1-0.05)^{\alpha+\beta-1}} \frac{\Gamma(\alpha+\beta)}{\Gamma(\alpha)\Gamma(\beta)} \quad \text{for all } x \in [0.05,\ 1],$$

where $\alpha$ and $\beta$ are the two shape parameters. We included the mean generation interval as a constant value [8] into our model and determined the probability that the viral genome of a confirmed case gets sequenced by dividing the monthly number of sequences by the monthly number of confirmed cases.

Supplementary tables S7A and S7B present an overview of the prior distributions as well as the fixed parameters.

| Symbol | Comment | Support | Prior |
|---|---|---|---|
| $R_e$ | Effective reproduction number | $]0, \infty[$ | Gamma(shape = 10, rate = 10) |
| $k$ | Dispersion parameter | $]0, \infty[$ | Gamma(shape = 5, rate = 10) |
| $\mu$ | Mutation probability | $[0, 1]$ | Beta(shape = 27, shape = 68) |
| $M$ | Yearly mutation rate | $]-\infty, \infty[$ | Normal(mean = 14, variance = 0.25) |
| $\tau_{\text{test}}$ | Testing probability | $[0.05, 1]$ | ScaledBeta(shape = 1, shape = 3) |

**Table S7A. Summary of parameters of prior distributions.**

| Symbol | Comment | Value | Unit | Source |
|---|---|---|---|---|
| $D$ | Mean generation interval | 5.2 | days | Ganyani et al. 2020 |
| $\tau_{\text{sequence}}$ | Sequencing probability | 1.42 − 85.94% | percentage | see column Nos/Nocc of tables S1, S3 and S5 |

**Table S7B. Summary of fixed parameters.**

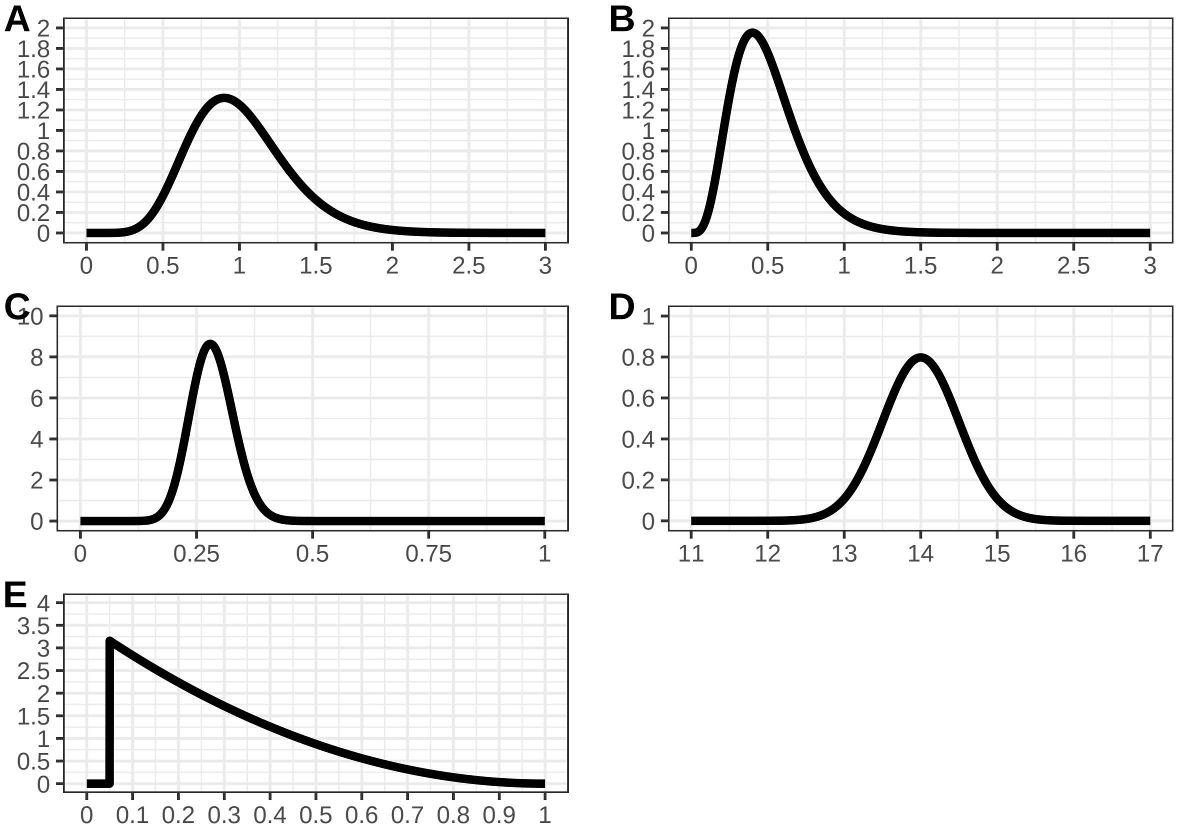

**Figure S1. Prior distributions.** A: Prior distribution of the effective reproduction number $R_e$. B: Prior distribution of the dispersion parameter $k$. C: Prior distribution of the mutation probability $\mu$. D: Prior distribution of the yearly mutation rate $M$. E: Prior distribution of the testing probability $\tau_{test}$.

### 5 Simulations

In this chapter, we present three alternatives to Figure 2 to display the results of the simulation study. We use two different methods to assess the precision of the parameter estimates. First, we use the coefficient of variation as error metric between the true value and the estimated mean of the posterior distribution. Second, we measure the error between the true value and the estimated mean of the posterior distribution applying the root mean square error (RMSE). Third, for each combination of input values we assess the proportion of repetitions of the parameter estimate in which the true value is contained in the estimated 95% credible interval of the posterior distribution.

#### 5.1 Coefficient of variation

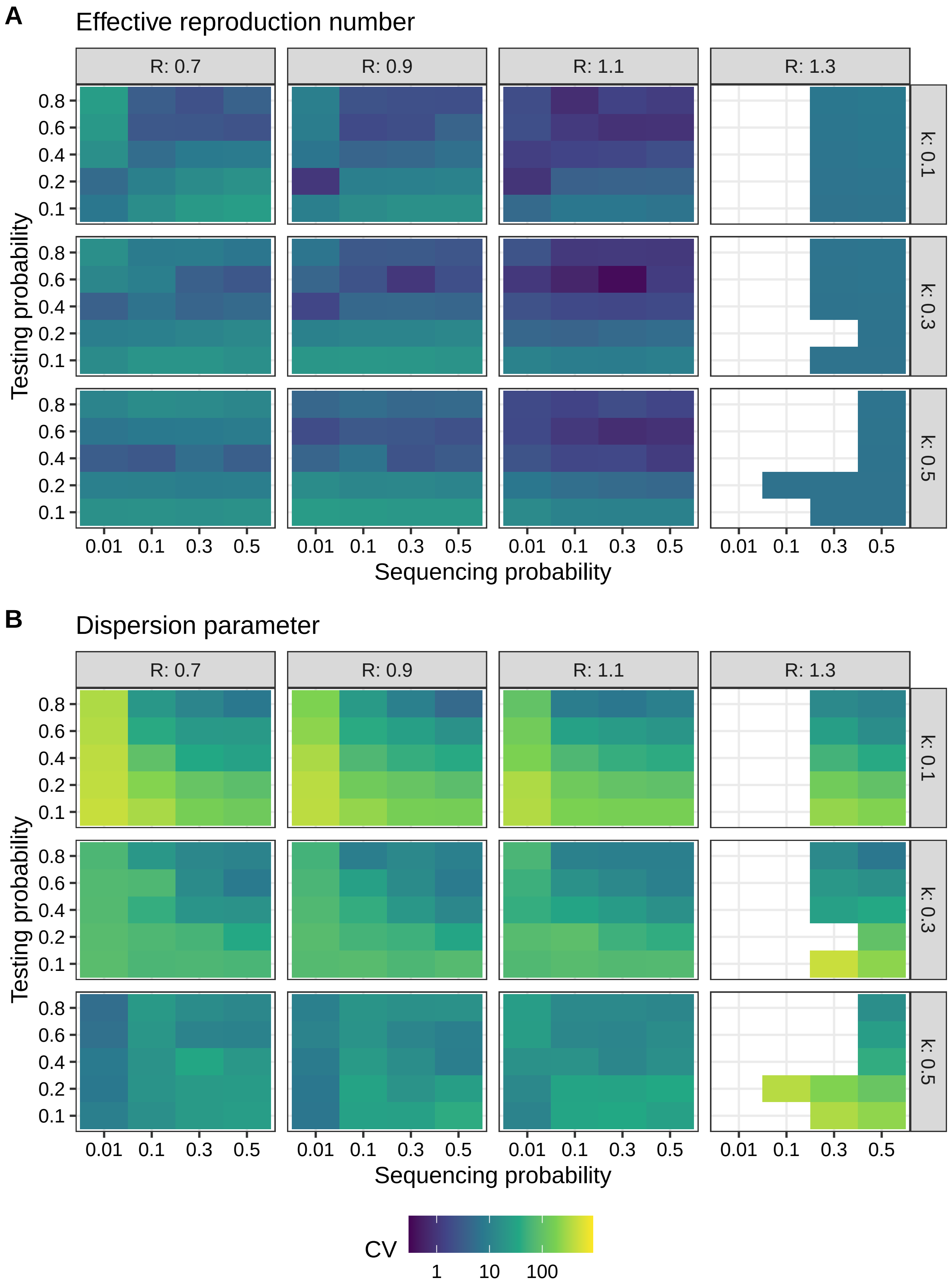

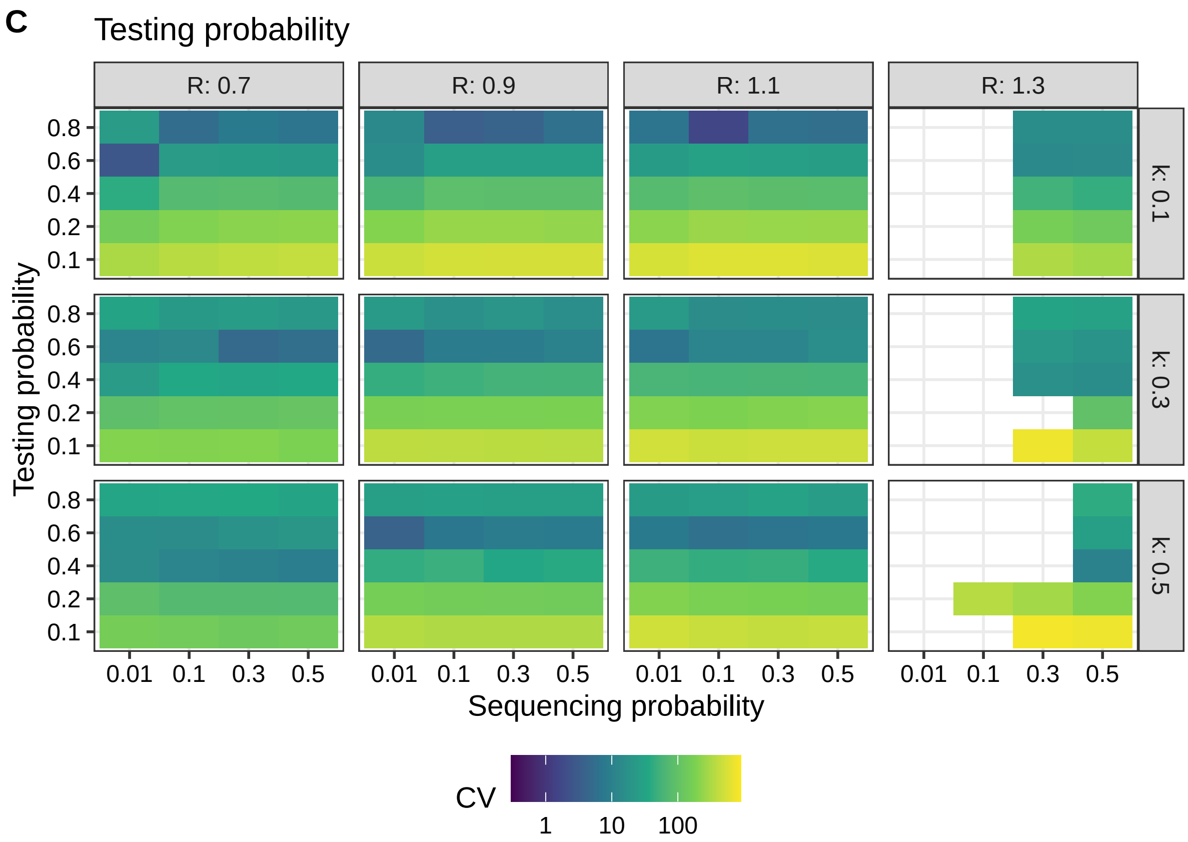

**Figure S2. Validation of the Bayesian inference model to estimate transmission parameters from the size distribution of identical sequence clusters.** A: Coefficient of variation of the estimate of the effective reproduction number $R_e$. B: Coefficient of variation of the estimate of the dispersion parameter $\tilde{k}$. C: Coefficient of variation of the estimate of the testing probability $\tilde{\tau}_{test}$. The coefficient of variation is computed as 100 times the ratio of the root mean squared error (RMSE) between the true value and the estimated mean of the posterior distribution to the true value. For each parameter combination, we ran the model 10 times on $3,000$ simulated clusters each.

#### 5.2 Root mean square error

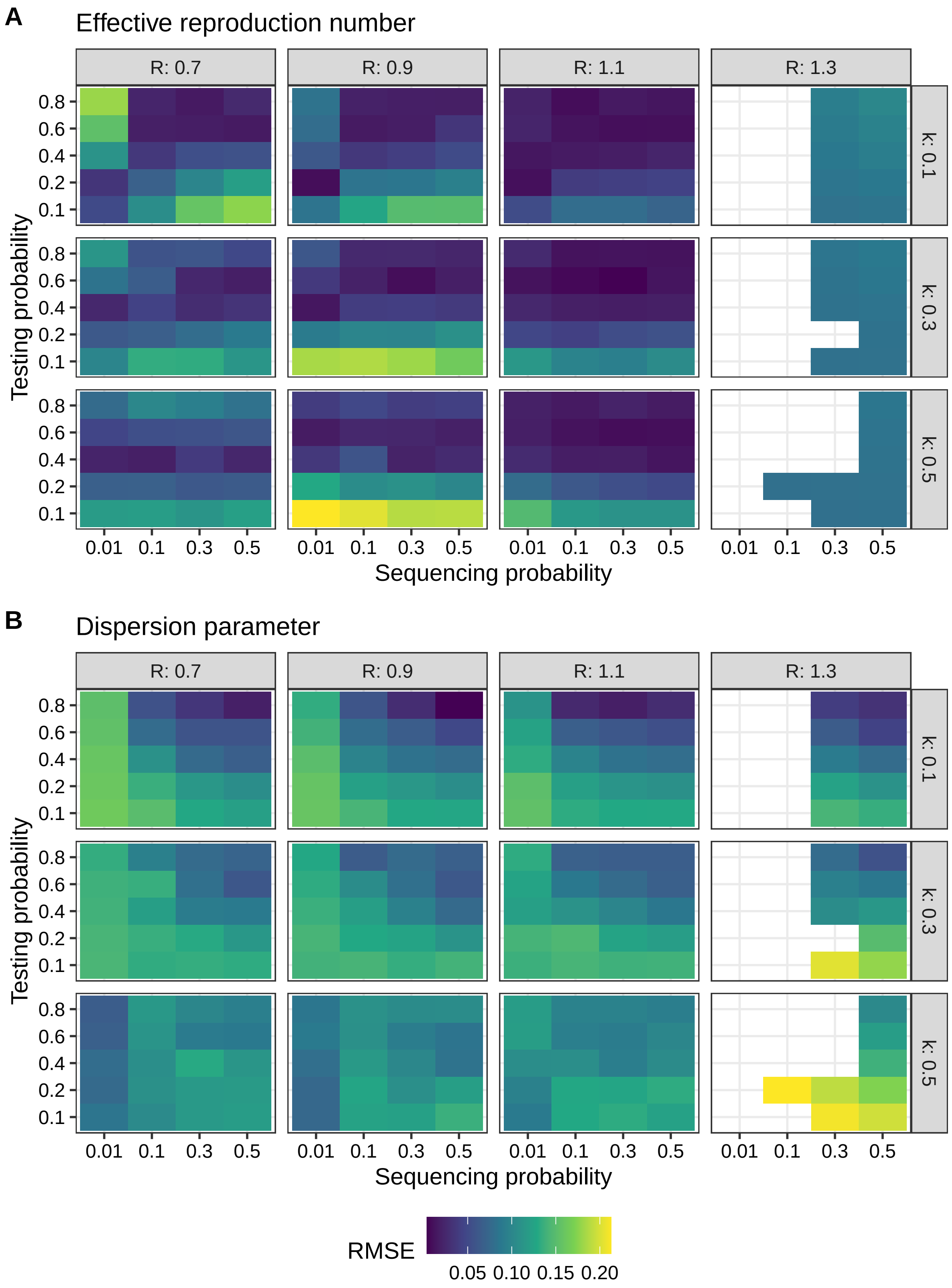

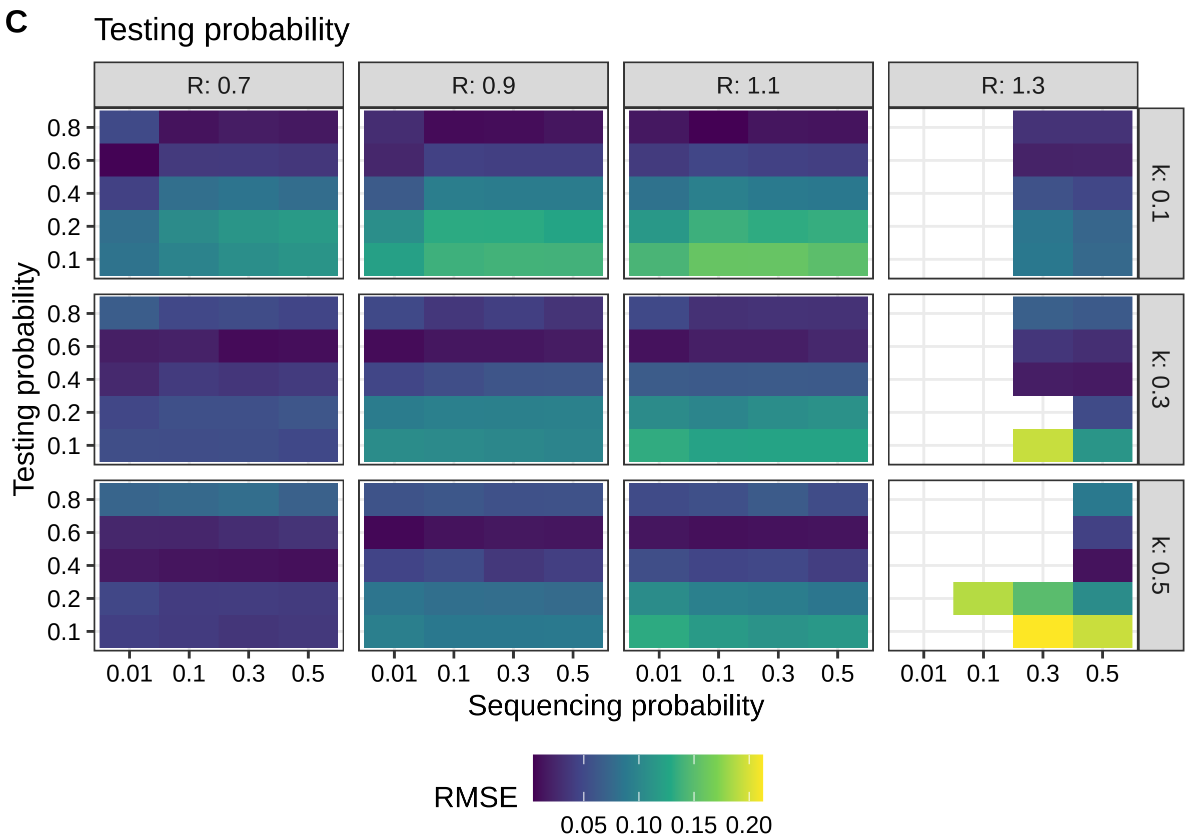

214

**Figure S3. Validation of the Bayesian inference model to estimate transmission parameters from the size distribution of identical sequence clusters.** A: Error in estimating the effective reproduction number $R_e$. B: Error in estimating the dispersion parameter˜$k$. C: Error in estimating the testing probability˜$\tau_{test}$. Error is measured by the root mean square error (RMSE) between the true value and the estimated mean of the posterior distribution. For each parameter combination, we ran the model 10 times on 3,000 simulated clusters each.

#### 5.3 Coverage

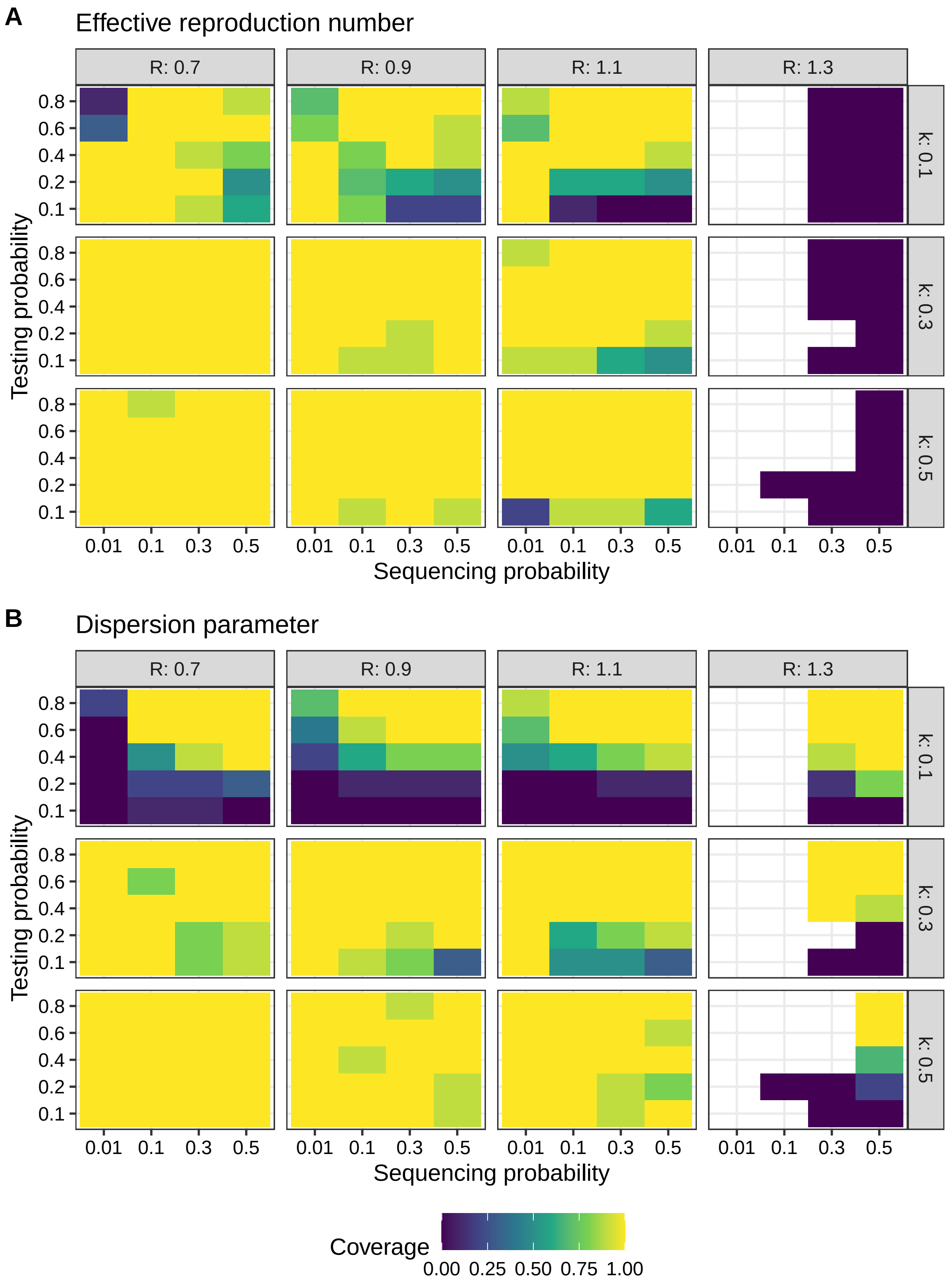
A
Effective reproduction number
R: 0.7
R: 0.9
R: 1.1
R: 1.3
k: 0.1
k: 0.3
k: 0.5
Testing probability
0.8
0.6
0.4
0.2
0.1
0.01
0.1
0.3
0.5
Sequencing probability
B
Dispersion parameter
Coverage
0.00
0.25
0.50
0.75
1.00

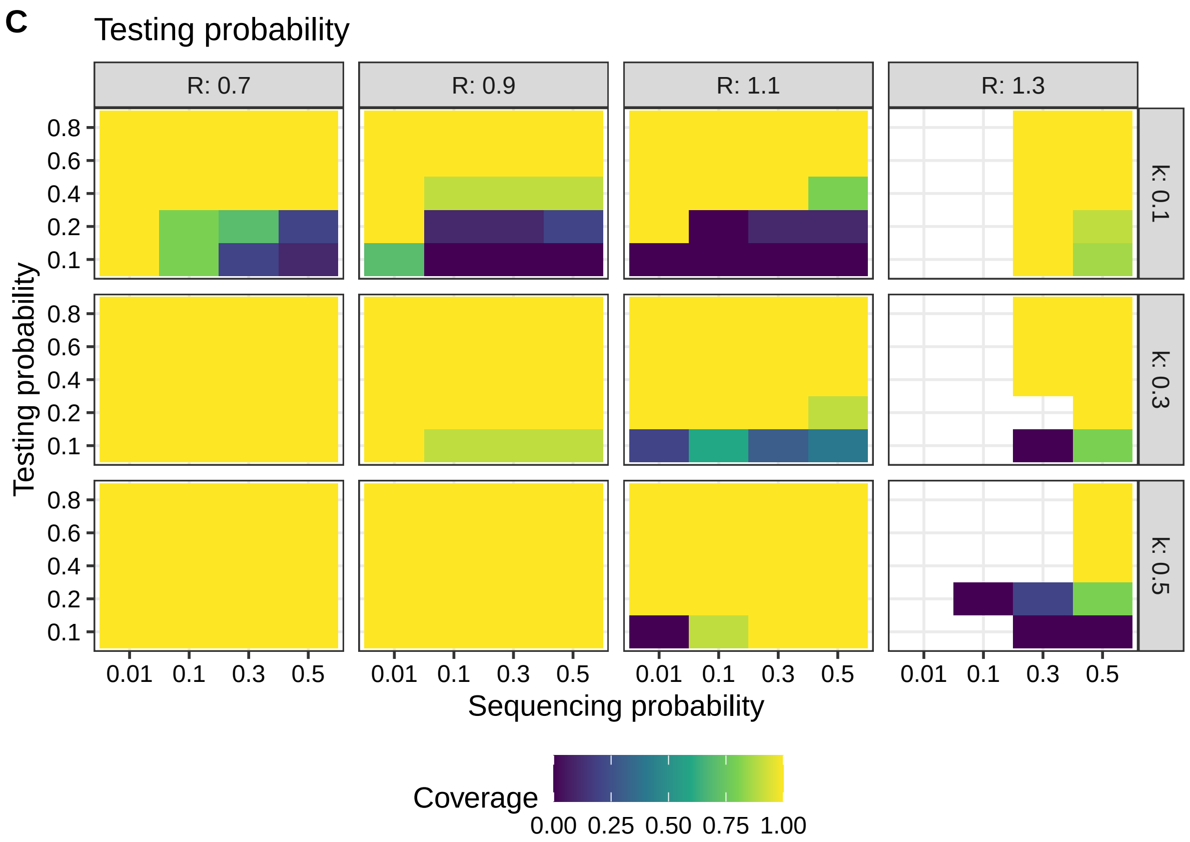

**Figure S4. Validation of the Bayesian inference model to estimate transmission parameters from the size distribution of identical sequence clusters.** A: Coverage of true value by 95% credible interval of the posterior distribution of the effective reproduction number $R_e$. B: Coverage of true value by 95% credible interval of the posterior distribution of the effective reproduction number $k$. C: Coverage of true value by 95% credible interval of the posterior distribution of the effective reproduction number $M$. D: Coverage of true value by 95% credible interval of the posterior distribution of the effective reproduction number $\tau_{test}$. For each parameter combination, we ran the model 10 times on 3000 simulated clusters each.

### 6 Additional results

In the following tables we present the parameter estimates based on the size distribution of identical SARS-CoV-2 sequence clusters in Switzerland, Denmark and Germany in 2021 in numerical form. The results for the effective reproduction number $R_e$, the dispersion parameter $k$ and the testing probability $\tau_{test}$ are also contained in Figure 4 of the manuscript. For each country, month and parameter, estimates are summarized by mean and 95% credible interval of the posterior distribution.

#### 6.1 Switzerland

| Month | Effective reproduction number | Dispersion parameter |
|---|---|---|
| January | 0.902 [0.797-1.035] | 0.081 [0.059-0.101] |
| February | 1.114 [0.989-1.275] | 0.252 [0.156-0.355] |
| March | 1.095 [0.973-1.254] | 0.191 [0.123-0.256] |
| April | 1.053 [0.932-1.212] | 0.232 [0.157-0.303] |
| May | 1.049 [0.919-1.217] | 0.248 [0.158-0.336] |
| June | 1.100 [0.975-1.254] | 0.183 [0.107-0.259] |
| July | 0.978 [0.863-1.115] | 0.195 [0.159-0.236] |
| August | 0.901 [0.793-1.036] | 0.129 [0.109-0.149] |
| September | 1.005 [0.890-1.144] | 0.156 [0.126-0.184] |
| October | 1.023 [0.907-1.174] | 0.212 [0.165-0.255] |
| November | 1.017 [0.911-1.163] | 0.115 [0.090-0.138] |
| December | 1.003 [0.892-1.137] | 0.024 [0.017-0.030] |

**Table S8A. Results of parameter estimations (Switzerland).** Samples of posterior distributions of the effective reproduction number and the dispersion parameter summarized by mean and 95% credible interval.

| Month | Mutation probability | Testing probability | Detection probability |
|---|---|---|---|
| January | 0.288 [0.203-0.377] | 0.869 [0.575-0.995] | 0.096 [0.063-0.110] |
| February | 0.281 [0.201-0.370] | 0.736 [0.297-0.992] | 0.097 [0.039-0.130] |
| March | 0.285 [0.203-0.373] | 0.788 [0.382-0.992] | 0.092 [0.045-0.116] |
| April | 0.279 [0.196-0.370] | 0.781 [0.372-0.991] | 0.101 [0.048-0.128] |
| May | 0.283 [0.194-0.381] | 0.731 [0.285-0.987] | 0.124 [0.049-0.168] |
| June | 0.280 [0.199-0.368] | 0.779 [0.340-0.992] | 0.167 [0.073-0.213] |
| July | 0.283 [0.195-0.368] | 0.885 [0.637-0.997] | 0.299 [0.215-0.337] |
| August | 0.289 [0.201-0.380] | 0.924 [0.724-0.998] | 0.178 [0.139-0.192] |
| September | 0.284 [0.198-0.372] | 0.899 [0.653-0.997] | 0.141 [0.102-0.156] |
| October | 0.283 [0.199-0.376] | 0.835 [0.509-0.994] | 0.213 [0.130-0.253] |
| November | 0.283 [0.205-0.373] | 0.882 [0.610-0.996] | 0.086 [0.059-0.097] |
| December | 0.283 [0.202-0.370] | 0.884 [0.603-0.997] | 0.029 [0.020-0.033] |

**Table S8B. Results of parameter estimations (Switzerland).** Samples of posterior distributions of the mutation probability, the testing probability and the detection probability summarized by mean and 95% credible interval.

#### 6.2 Denmark

| Month | Effective reproduction number | Dispersion parameter |
|---|---|---|
| January | 1.228 [1.093-1.391] | 0.400 [0.310-0.478] |
| February | 1.282 [1.133-1.471] | 0.556 [0.401-0.695] |
| March | 1.214 [1.085-1.373] | 0.512 [0.423-0.591] |
| April | 1.248 [1.112-1.419] | 0.522 [0.430-0.611] |
| May | 1.184 [1.052-1.351] | 0.351 [0.313-0.390] |
| June | 1.294 [1.157-1.461] | 0.343 [0.247-0.434] |
| July | 1.058 [0.937-1.207] | 0.281 [0.254-0.310] |
| August | 1.017 [0.904-1.170] | 0.420 [0.385-0.457] |
| September | 1.125 [1.000-1.284] | 0.407 [0.344-0.465] |
| October | 1.180 [1.052-1.345] | 0.402 [0.361-0.445] |
| November | 1.131 [1.005-1.285] | 0.248 [0.229-0.265] |
| December | 1.218 [1.094-1.379] | 0.134 [0.117-0.149] |

**Table S9A. Results of parameter estimations (Denmark).** Samples of posterior distributions of the effective reproduction number and the dispersion parameter summarized by mean and 95% credible interval.

| Month | Mutation probability | Testing probability | Detection probability |
|---|---|---|---|
| January | 0.278 [0.198-0.367] | 0.757 [0.316-0.992] | 0.349 [0.146-0.457] |
| February | 0.278 [0.189-0.370] | 0.630 [0.146-0.984] | 0.454 [0.105-0.709] |
| March | 0.278 [0.199-0.365] | 0.778 [0.363-0.993] | 0.657 [0.306-0.839] |
| April | 0.278 [0.197-0.365] | 0.785 [0.345-0.993] | 0.562 [0.247-0.711] |
| May | 0.278 [0.195-0.371] | 0.892 [0.618-0.997] | 0.695 [0.481-0.777] |
| June | 0.274 [0.193-0.359] | 0.748 [0.317-0.993] | 0.326 [0.138-0.433] |
| July | 0.283 [0.198-0.370] | 0.946 [0.806-0.998] | 0.786 [0.670-0.830] |
| August | 0.280 [0.198-0.373] | 0.927 [0.747-0.998] | 0.762 [0.614-0.820] |
| September | 0.282 [0.201-0.373] | 0.837 [0.498-0.995] | 0.719 [0.428-0.855] |
| October | 0.281 [0.200-0.369] | 0.904 [0.684-0.998] | 0.709 [0.536-0.783] |
| November | 0.280 [0.191-0.372] | 0.953 [0.830-0.998] | 0.443 [0.386-0.464] |
| December | 0.279 [0.200-0.366] | 0.941 [0.787-0.999] | 0.110 [0.092-0.117] |

**Table S9B. Results of parameter estimations (Denmark).** Samples of posterior distributions of the mutation probability, the testing probability and the detection probability summarized by mean and 95% credible interval.

#### 6.3 Germany

| Month | Effective reproduction number | Dispersion parameter |
|---|---|---|
| January | 1.095 [0.979-1.243] | 0.020 [0.013-0.027] |
| February | 1.096 [0.973-1.257] | 0.114 [0.090-0.133] |
| March | 1.125 [1.008-1.270] | 0.182 [0.150-0.208] |
| April | 1.124 [1.005-1.295] | 0.179 [0.148-0.203] |
| May | 1.110 [0.997-1.242] | 0.197 [0.164-0.226] |
| June | 1.084 [0.960-1.234] | 0.253 [0.190-0.313] |
| July | 1.023 [0.912-1.169] | 0.223 [0.176-0.267] |
| August | 0.965 [0.854-1.111] | 0.186 [0.165-0.206] |
| September | 0.986 [0.877-1.141] | 0.208 [0.188-0.230] |
| October | 1.063 [0.947-1.214] | 0.137 [0.117-0.156] |
| November | 1.036 [0.922-1.186] | 0.080 [0.069-0.090] |
| December | 0.962 [0.855-1.098] | 0.034 [0.031-0.037] |

**Table S10A. Results of parameter estimations (Germany).** Samples of posterior distributions of the effective reproduction number and the dispersion parameter summarized by mean and 95% credible interval.

| Month | Mutation probability | Testing probability | Detection probability |
|---|---|---|---|
| January | 0.282 [0.202-0.368] | 0.859 [0.554-0.996] | 0.012 [0.008-0.014] |
| February | 0.283 [0.197-0.378] | 0.901 [0.662-0.997] | 0.073 [0.054-0.081] |
| March | 0.280 [0.198-0.366] | 0.896 [0.641-0.998] | 0.082 [0.059-0.092] |
| April | 0.279 [0.198-0.374] | 0.898 [0.645-0.998] | 0.066 [0.047-0.073] |
| May | 0.281 [0.203-0.359] | 0.897 [0.649-0.998] | 0.081 [0.059-0.091] |
| June | 0.281 [0.197-0.370] | 0.836 [0.482-0.995] | 0.141 [0.081-0.168] |
| July | 0.282 [0.203-0.374] | 0.870 [0.560-0.997] | 0.182 [0.117-0.208] |
| August | 0.286 [0.197-0.379] | 0.937 [0.763-0.998] | 0.155 [0.126-0.165] |
| September | 0.283 [0.200-0.382] | 0.941 [0.796-0.998] | 0.135 [0.114-0.143] |
| October | 0.283 [0.198-0.371] | 0.919 [0.728-0.997] | 0.076 [0.061-0.083] |
| November | 0.282 [0.195-0.375] | 0.939 [0.776-0.998] | 0.035 [0.029-0.038] |
| December | 0.288 [0.203-0.378] | 0.964 [0.878-0.999] | 0.038 [0.034-0.039] |

**Table S10B. Results of parameter estimations (Germany).** Samples of posterior distributions of the mutation probability, the testing probability and the detection probability summarized by mean and 95% credible interval.

### 7 Sensitivity analysis

We developed several modifications of our inference model to assess our modelling assumptions regarding the mutation and the case detection process. In the following we present the results of the parameter estimations when we use a prior distribution for the yearly mutation rate $M$ instead of a prior distribution for the mutation probability $\mu$ to model the mutation process. Furthermore, we also reran our model using constant values for the testing probability instead of a prior distribution.

As an alternative to using a prior distribution that incorporates uncertainty for the testing probability in our model, we used estimates of the ascertainment rate from [11] as constant values. We set the testing probability for the months of January to August to 57.6% and for the months of September to December to 35%.

#### 7.1 Prior distribution for mutation probability and constant testing probability

We use a beta distribution with shape parameters $\alpha = 27$ and $\beta = 68$ as prior distribution for the mutation probability constant values for the testing probability (57.6% for the months of January to August and 35% for the months of September to December). The following figures and tables present the results.

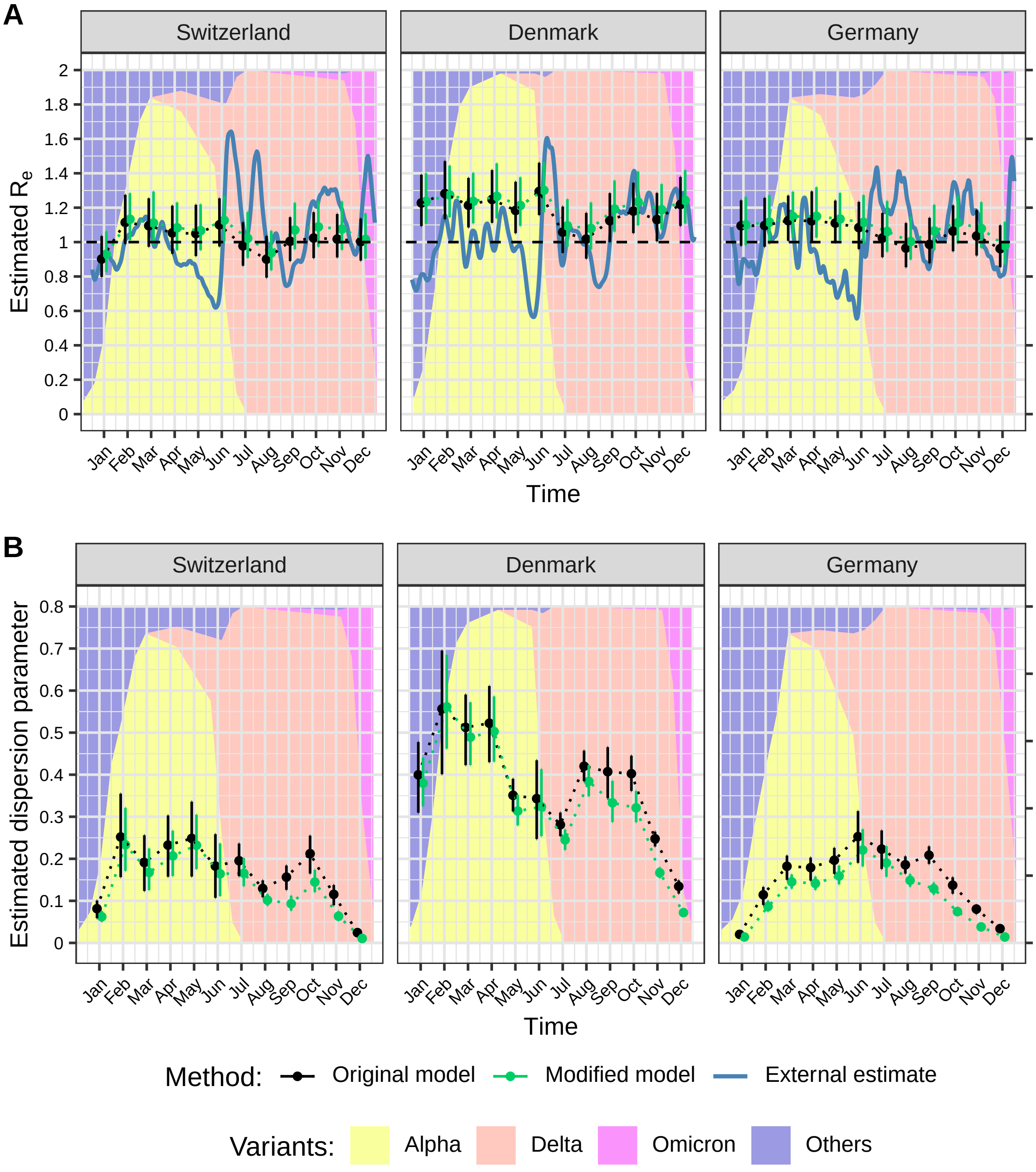

**Figure S5. Comparison of the estimated effective reproduction numbers and dispersion parameters by country and month.** A: Effective reproduction number $R_e$. B: Dispersion parameter $k$. The estimates are based on monthly time windows of identical sequence clusters. For each month the estimated means and 95% credible intervals of the posterior distribution obtained by two different models are shown. $R_e$ values are compared to external estimates based on laboratory-confirmed cases (in blue, from github.com/covid-19-Re [12]).

##### 7.1.1 Switzerland

| Month | Effective reproduction number | Dispersion parameter |
|---|---|---|
| January | 0.930 [0.826-1.062] | 0.062 [0.050-0.076] |
| February | 1.133 [1.011-1.284] | 0.233 [0.171-0.321] |
| March | 1.111 [0.984-1.294] | 0.167 [0.126-0.224] |
| April | 1.083 [0.953-1.230] | 0.207 [0.159-0.267] |
| May | 1.067 [0.953-1.220] | 0.232 [0.176-0.305] |
| June | 1.127 [1.003-1.268] | 0.164 [0.111-0.236] |
| July | 1.022 [0.908-1.174] | 0.166 [0.135-0.202] |
| August | 0.938 [0.836-1.066] | 0.102 [0.089-0.116] |
| September | 1.070 [0.959-1.228] | 0.093 [0.077-0.112] |
| October | 1.089 [0.972-1.238] | 0.145 [0.121-0.174] |
| November | 1.075 [0.955-1.236] | 0.063 [0.053-0.076] |
| December | 1.015 [0.904-1.167] | 0.011 [0.009-0.012] |

**Table S11A. Results of parameter estimations with constant testing probability (Switzerland).** Samples of posterior distributions of the effective reproduction number and the dispersion parameter summarized by mean and 95% credible interval.

| Month | Mutation probability | Testing probability | Detection probability |
|---|---|---|---|
| January | 0.289 [0.203-0.379] | 0.576 | 0.063 |
| February | 0.282 [0.198-0.368] | 0.576 | 0.076 |
| March | 0.280 [0.190-0.383] | 0.576 | 0.067 |
| April | 0.282 [0.190-0.370] | 0.576 | 0.074 |
| May | 0.281 [0.200-0.371] | 0.576 | 0.098 |
| June | 0.284 [0.204-0.366] | 0.576 | 0.124 |
| July | 0.283 [0.201-0.378] | 0.576 | 0.195 |
| August | 0.284 [0.198-0.373] | 0.576 | 0.111 |
| September | 0.283 [0.202-0.378] | 0.350 | 0.055 |
| October | 0.279 [0.193-0.367] | 0.350 | 0.089 |
| November | 0.286 [0.201-0.379] | 0.350 | 0.034 |
| December | 0.282 [0.202-0.372] | 0.350 | 0.012 |

**Table S11B. Results of parameter estimations with constant testing probability (Switzerland).** Samples of posterior distributions of the mutation probability summarized by mean and 95% credible interval and constant input values of the testing probability and the detection probability.

##### 7.1.2 Denmark

| Month | Effective reproduction number | Dispersion parameter |
|---|---|---|
| January | 1.237 [1.107-1.401] | 0.380 [0.325-0.440] |
| February | 1.274 [1.144-1.445] | 0.561 [0.462-0.685] |
| March | 1.236 [1.108-1.403] | 0.489 [0.423-0.572] |
| April | 1.266 [1.138-1.458] | 0.503 [0.431-0.586] |
| May | 1.213 [1.094-1.377] | 0.314 [0.279-0.352] |
| June | 1.301 [1.173-1.464] | 0.324 [0.254-0.413] |
| July | 1.094 [0.973-1.249] | 0.245 [0.221-0.270] |
| August | 1.079 [0.957-1.230] | 0.384 [0.349-0.423] |
| September | 1.192 [1.060-1.358] | 0.333 [0.287-0.385] |
| October | 1.233 [1.098-1.410] | 0.321 [0.287-0.361] |
| November | 1.188 [1.073-1.337] | 0.167 [0.154-0.180] |
| December | 1.242 [1.108-1.418] | 0.072 [0.065-0.081] |

**Table S12A. Results of parameter estimations with constant testing probability (Denmark).** Samples of posterior distributions of the effective reproduction number and the dispersion parameter summarized by mean and 95% credible interval.

| Month | Mutation probability | Testing probability | Detection probability |
|---|---|---|---|
| January | 0.275 [0.194-0.362] | 0.576 | 0.266 |
| February | 0.273 [0.192-0.362] | 0.576 | 0.415 |
| March | 0.280 [0.201-0.365] | 0.576 | 0.487 |
| April | 0.279 [0.203-0.376] | 0.576 | 0.412 |
| May | 0.278 [0.201-0.366] | 0.576 | 0.449 |
| June | 0.274 [0.198-0.358] | 0.576 | 0.251 |
| July | 0.276 [0.190-0.366] | 0.576 | 0.479 |
| August | 0.283 [0.195-0.377] | 0.576 | 0.473 |
| September | 0.279 [0.193-0.370] | 0.350 | 0.301 |
| October | 0.275 [0.189-0.368] | 0.350 | 0.274 |
| November | 0.276 [0.200-0.358] | 0.350 | 0.163 |
| December | 0.275 [0.192-0.366] | 0.350 | 0.041 |

**Table S12B. Results of parameter estimations with constant testing probability (Denmark).** Samples of posterior distributions of the mutation probability summarized by mean and 95% credible interval and constant input values of the testing probability and the detection probability.

##### 7.1.3 Germany

| Month | Effective reproduction number | Dispersion parameter |
|---|---|---|
| January | 1.100 [0.982-1.262] | 0.014 [0.011-0.019] |
| February | 1.115 [0.989-1.273] | 0.087 [0.074-0.100] |
| March | 1.149 [1.024-1.293] | 0.145 [0.129-0.163] |
| April | 1.150 [1.026-1.319] | 0.141 [0.126-0.158] |
| May | 1.134 [1.010-1.286] | 0.159 [0.139-0.184] |
| June | 1.115 [0.993-1.273] | 0.221 [0.182-0.270] |
| July | 1.061 [0.944-1.239] | 0.190 [0.157-0.230] |
| August | 1.003 [0.898-1.147] | 0.149 [0.134-0.164] |
| September | 1.065 [0.953-1.217] | 0.129 [0.115-0.144] |
| October | 1.113 [0.993-1.268] | 0.074 [0.066-0.084] |
| November | 1.079 [0.966-1.232] | 0.038 [0.034-0.043] |
| December | 0.979 [0.872-1.117] | 0.014 [0.013-0.015] |

**Table S13A. Results of parameter estimations with constant testing probability (Germany).** Samples of posterior distributions of the effective reproduction number and the dispersion parameter summarized by mean and 95% credible interval.

| Month | Mutation probability | Testing probability | Detection probability |
|---|---|---|---|
| January | 0.282 [0.198-0.374] | 0.576 | 0.008 |
| February | 0.280 [0.193-0.373] | 0.576 | 0.047 |
| March | 0.278 [0.194-0.361] | 0.576 | 0.053 |
| April | 0.279 [0.194-0.374] | 0.576 | 0.042 |
| May | 0.277 [0.191-0.366] | 0.576 | 0.052 |
| June | 0.281 [0.197-0.375] | 0.576 | 0.097 |
| July | 0.282 [0.197-0.385] | 0.576 | 0.120 |
| August | 0.279 [0.198-0.371] | 0.576 | 0.095 |
| September | 0.279 [0.195-0.370] | 0.350 | 0.050 |
| October | 0.280 [0.198-0.372] | 0.350 | 0.029 |
| November | 0.285 [0.204-0.375] | 0.350 | 0.013 |
| December | 0.283 [0.198-0.374] | 0.350 | 0.014 |

**Table S13B. Results of parameter estimations with constant testing probability (Germany).** Samples of posterior distributions of the mutation probability summarized by mean and 95% credible interval and constant input values of the testing probability and the detection probability.

#### 7.2 Constant mutation probability and prior distribution for testing probability

We use a constant mutation probability $\mu = 0.281$ and the same prior distribution for the testing probability as in the original model. The following figures and tables present the results.

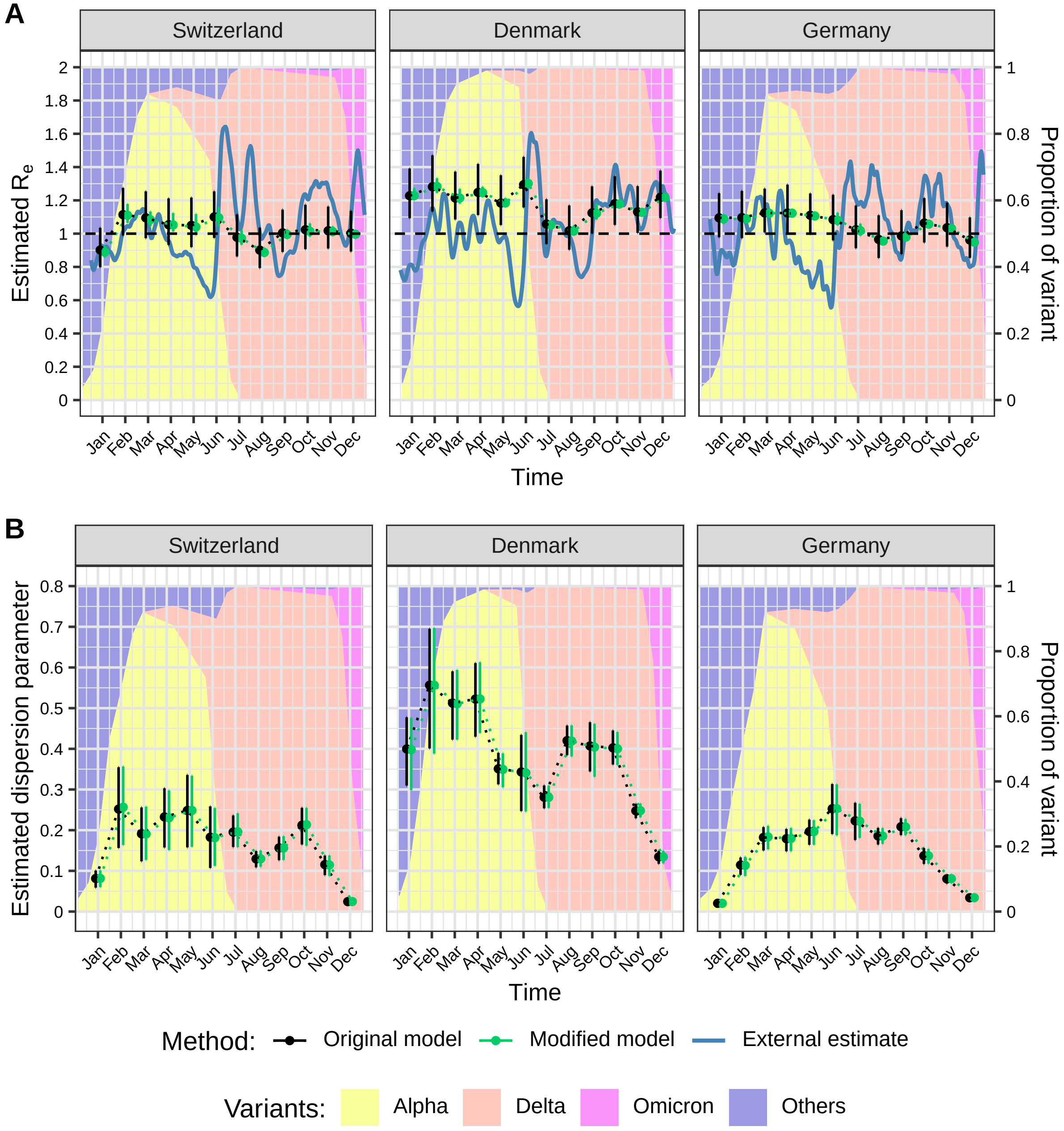

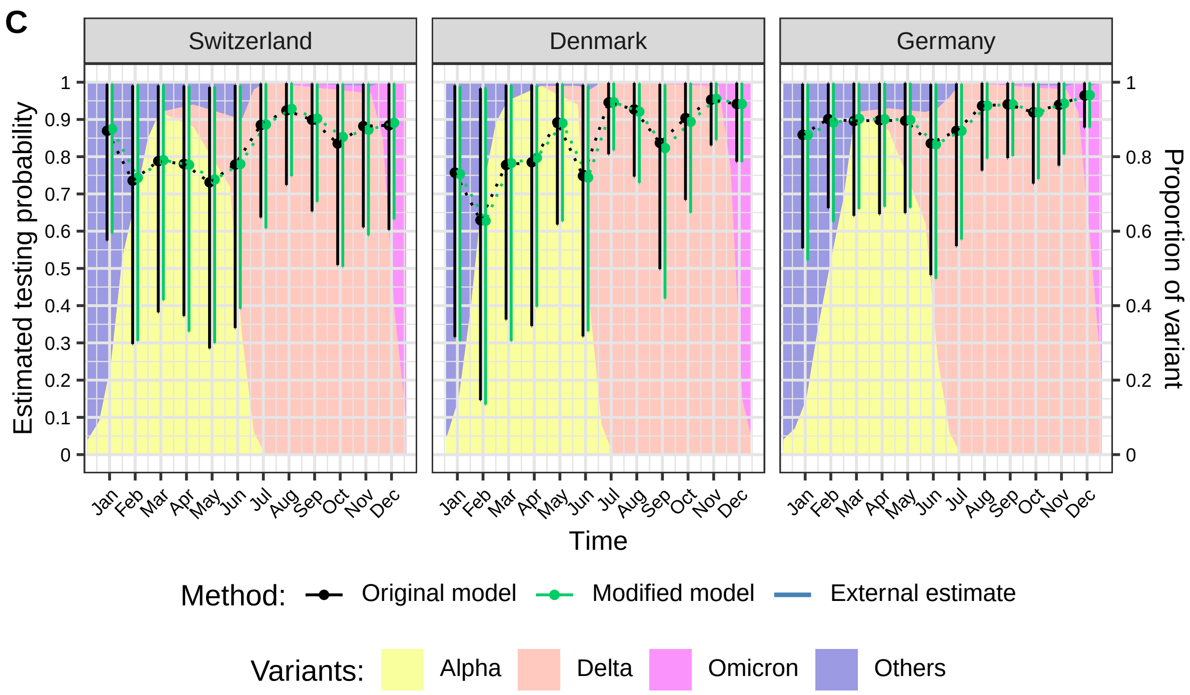

**Figure S6. Comparison of the estimated effective reproduction numbers, dispersion parameters and testing probabilities by country and month.** A: Effective reproduction number $R_e$. B: Dispersion parameter $k$. C: Testing probability $\tau_{test}$. The estimates are based on monthly time windows of identical sequence clusters. For each month the estimated means and 95% credible intervals of the posterior distribution obtained by two different models are shown. $R_e$ values are compared to external estimates based on laboratory-confirmed cases (in blue, from github.com/covid-19-Re [12]).

##### 7.2.1 Switzerland

| Month | Effective reproduction number | Dispersion parameter |
|---|---|---|
| January | 0.889 [0.857-0.925] | 0.081 [0.060-0.101] |
| February | 1.110 [1.065-1.178] | 0.256 [0.163-0.357] |
| March | 1.084 [1.045-1.133] | 0.191 [0.127-0.258] |
| April | 1.052 [1.012-1.124] | 0.230 [0.151-0.298] |
| May | 1.040 [0.992-1.118] | 0.249 [0.159-0.334] |
| June | 1.096 [1.041-1.160] | 0.182 [0.112-0.255] |
| July | 0.972 [0.932-1.021] | 0.196 [0.158-0.242] |
| August | 0.887 [0.863-0.916] | 0.130 [0.110-0.150] |
| September | 0.995 [0.971-1.025] | 0.156 [0.127-0.185] |
| October | 1.014 [0.982-1.062] | 0.214 [0.162-0.255] |
| November | 1.012 [0.989-1.044] | 0.114 [0.088-0.138] |
| December | 0.996 [0.971-1.019] | 0.025 [0.018-0.030] |

**Table S14A. Results of parameter estimations with constant mutation probability (Switzerland).** Samples of posterior distributions of the effective reproduction number and the dispersion parameter summarized by mean and 95% credible interval.

| Month | Mutation probability | Testing probability | Detection probability |
|---|---|---|---|
| January | 0.281 | 0.874 [0.594-0.996] | 0.096 [0.066-0.110] |
| February | 0.281 | 0.743 [0.305-0.993] | 0.098 [0.040-0.130] |
| March | 0.281 | 0.791 [0.414-0.994] | 0.092 [0.048-0.116] |
| April | 0.281 | 0.778 [0.330-0.990] | 0.100 [0.043-0.127] |
| May | 0.281 | 0.738 [0.299-0.989] | 0.126 [0.051-0.168] |
| June | 0.281 | 0.780 [0.392-0.991] | 0.168 [0.084-0.213] |
| July | 0.281 | 0.887 [0.608-0.997] | 0.300 [0.206-0.337] |
| August | 0.281 | 0.928 [0.748-0.998] | 0.179 [0.144-0.192] |
| September | 0.281 | 0.902 [0.679-0.995] | 0.141 [0.106-0.156] |
| October | 0.281 | 0.853 [0.504-0.996] | 0.217 [0.129-0.254] |
| November | 0.281 | 0.872 [0.589-0.996] | 0.085 [0.057-0.097] |
| December | 0.281 | 0.891 [0.632-0.997] | 0.029 [0.021-0.033] |

**Table S14B. Results of parameter estimations with constant mutation probability (Switzerland).** Constant input values of the mutation probability and samples of posterior distributions of the testing probability and the detection probability summarized by mean and 95% credible interval.

##### 7.2.2 Denmark

| Month | Effective reproduction number | Dispersion parameter |
|---|---|---|
| January | 1.228 [1.200-1.274] | 0.398 [0.298-0.476] |
| February | 1.283 [1.245-1.333] | 0.557 [0.388-0.697] |
| March | 1.214 [1.181-1.269] | 0.511 [0.422-0.594] |
| April | 1.247 [1.221-1.282] | 0.523 [0.439-0.613] |
| May | 1.184 [1.160-1.214] | 0.349 [0.305-0.389] |
| June | 1.302 [1.275-1.332] | 0.341 [0.246-0.441] |
| July | 1.052 [1.025-1.078] | 0.281 [0.255-0.310] |
| August | 1.015 [0.992-1.048] | 0.419 [0.381-0.458] |
| September | 1.122 [1.085-1.180] | 0.404 [0.331-0.461] |
| October | 1.177 [1.155-1.206] | 0.400 [0.360-0.441] |
| November | 1.128 [1.115-1.142] | 0.248 [0.231-0.265] |
| December | 1.218 [1.208-1.228] | 0.134 [0.118-0.149] |

**Table S15A. Results of parameter estimations with constant mutation probability (Denmark).** Samples of posterior distributions of the effective reproduction number and the dispersion parameter summarized by mean and 95% credible interval.

| Month | Mutation probability | Testing probability | Detection probability |
|---|---|---|---|
| January | 0.281 | 0.753 [0.305-0.989] | 0.347 [0.141-0.456] |
| February | 0.281 | 0.628 [0.134-0.986] | 0.452 [0.097-0.711] |
| March | 0.281 | 0.783 [0.305-0.993] | 0.661 [0.258-0.838] |
| April | 0.281 | 0.797 [0.397-0.991] | 0.571 [0.284-0.710] |
| May | 0.281 | 0.890 [0.627-0.996] | 0.693 [0.488-0.776] |
| June | 0.281 | 0.744 [0.331-0.990] | 0.324 [0.144-0.432] |
| July | 0.281 | 0.946 [0.817-0.998] | 0.787 [0.679-0.830] |
| August | 0.281 | 0.921 [0.730-0.997] | 0.757 [0.600-0.819] |
| September | 0.281 | 0.823 [0.419-0.994] | 0.708 [0.360-0.854] |
| October | 0.281 | 0.894 [0.649-0.998] | 0.701 [0.509-0.782] |
| November | 0.281 | 0.955 [0.844-0.999] | 0.444 [0.393-0.464] |
| December | 0.281 | 0.941 [0.785-0.998] | 0.110 [0.092-0.117] |

**Table S15B. Results of parameter estimations with constant mutation probability (Denmark).** Constant input values of the mutation probability and samples of posterior distributions of the testing probability and the detection probability summarized by mean and 95% credible interval.

##### 7.2.3 Germany

| Month | Effective reproduction number | Dispersion parameter |
|---|---|---|
| January | 1.088 [1.058-1.116] | 0.020 [0.012-0.027] |
| February | 1.089 [1.070-1.113] | 0.113 [0.087-0.134] |
| March | 1.122 [1.109-1.144] | 0.183 [0.153-0.211] |
| April | 1.123 [1.109-1.144] | 0.180 [0.148-0.205] |
| May | 1.106 [1.090-1.130] | 0.197 [0.164-0.224] |
| June | 1.080 [1.049-1.126] | 0.253 [0.187-0.313] |
| July | 1.017 [0.987-1.060] | 0.222 [0.181-0.265] |
| August | 0.955 [0.940-0.977] | 0.185 [0.166-0.205] |
| September | 0.979 [0.965-0.997] | 0.208 [0.187-0.228] |
| October | 1.057 [1.043-1.078] | 0.137 [0.117-0.154] |
| November | 1.030 [1.018-1.044] | 0.081 [0.070-0.091] |
| December | 0.948 [0.937-0.959] | 0.034 [0.031-0.037] |

**Table S16A. Results of parameter estimations with constant mutation probability (Germany).** Samples of posterior distributions of the effective reproduction number and the dispersion parameter summarized by mean and 95% credible interval.

| Month | Mutation probability | Testing probability | Detection probability |
|---|---|---|---|
| January | 0.281 | 0.858 [0.522-0.995] | 0.012 [0.007-0.014] |
| February | 0.281 | 0.892 [0.625-0.998] | 0.073 [0.051-0.081] |
| March | 0.281 | 0.902 [0.660-0.997] | 0.083 [0.061-0.092] |
| April | 0.281 | 0.901 [0.666-0.998] | 0.066 [0.049-0.073] |
| May | 0.281 | 0.899 [0.663-0.998] | 0.082 [0.060-0.091] |
| June | 0.281 | 0.833 [0.472-0.996] | 0.141 [0.080-0.168] |
| July | 0.281 | 0.869 [0.578-0.996] | 0.182 [0.121-0.208] |
| August | 0.281 | 0.937 [0.795-0.999] | 0.155 [0.131-0.165] |
| September | 0.281 | 0.940 [0.802-0.998] | 0.135 [0.115-0.143] |
| October | 0.281 | 0.919 [0.740-0.997] | 0.076 [0.062-0.083] |
| November | 0.281 | 0.943 [0.806-0.999] | 0.035 [0.030-0.038] |
| December | 0.281 | 0.965 [0.878-0.999] | 0.038 [0.034-0.039] |

**Table S16B. Results of parameter estimations with constant mutation probability (Germany).** Constant input values of the mutation probability and samples of posterior distributions of the testing probability and the detection probability summarized by mean and 95% credible interval.

##### 7.3 Constant mutation probability and constant testing probability

We use a constant mutation probability $\mu = 28.1\%$ and constant values for the testing probability (57.6% for the months of January to August and 35% for the months of September to December). The following figures and tables present the results.

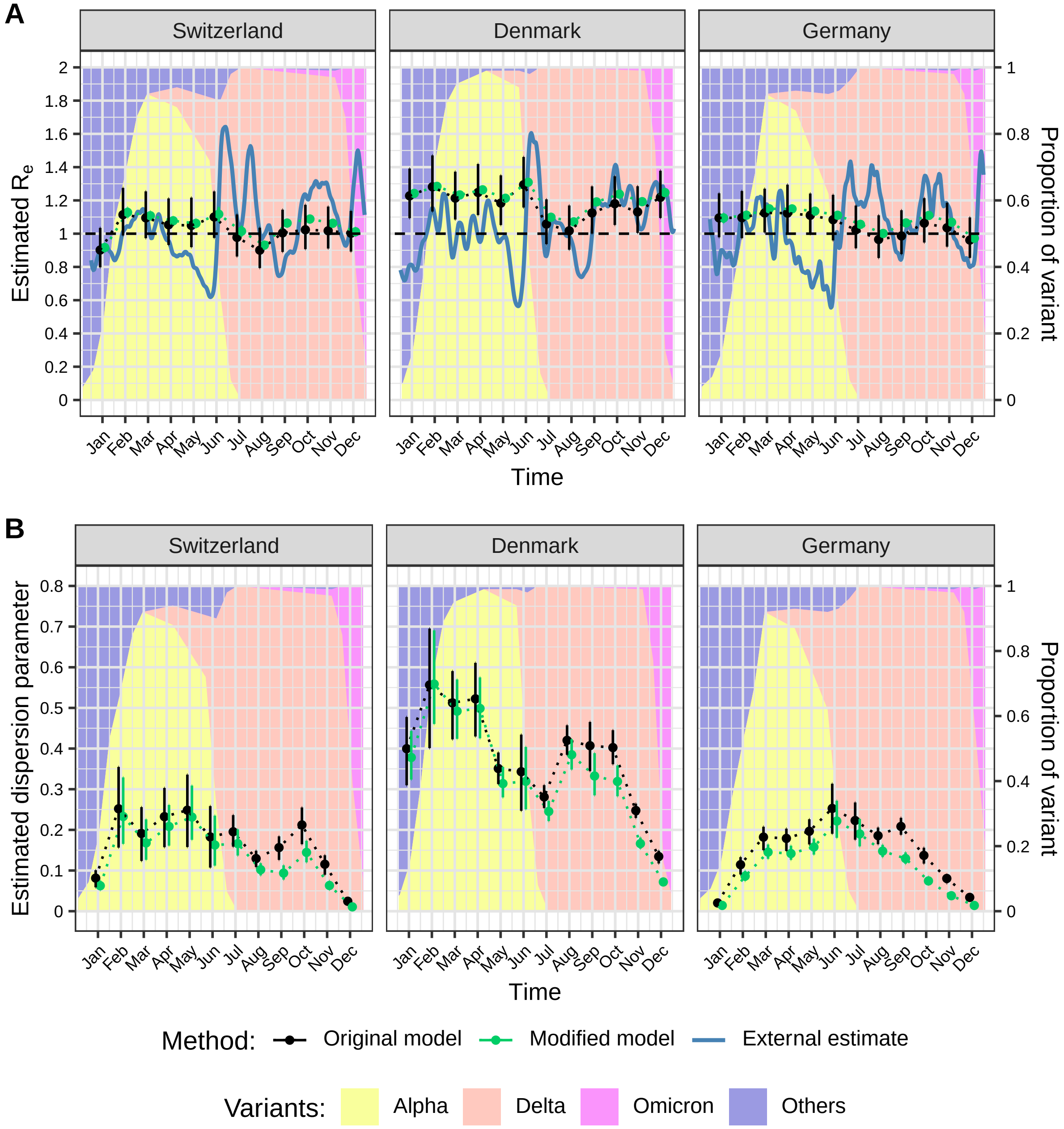

**Figure S7. Comparison of the estimated effective reproduction numbers and dispersion parameters by country and month.** A: Effective reproduction number $R_e$. B: Dispersion parameter $k$. The estimates are based on monthly time windows of identical sequence clusters. For each month the estimated means and 95% credible intervals of the posterior distribution obtained by two different models are shown. $R_e$ values are compared to external estimates based on laboratory-confirmed cases (in blue, from github.com/covid-19-Re [12]).

##### 7.3.1 Switzerland

| Month | Effective reproduction number | Dispersion parameter |
|---|---|---|
| January | 0.917 [0.888-0.942] | 0.063 [0.052-0.075] |
| February | 1.128 [1.097-1.157] | 0.233 [0.166-0.329] |
| March | 1.108 [1.082-1.134] | 0.168 [0.126-0.226] |
| April | 1.078 [1.054-1.100] | 0.208 [0.161-0.261] |
| May | 1.062 [1.036-1.087] | 0.232 [0.176-0.309] |
| June | 1.117 [1.073-1.159] | 0.163 [0.112-0.235] |
| July | 1.015 [0.983-1.047] | 0.166 [0.137-0.201] |
| August | 0.931 [0.911-0.952] | 0.102 [0.088-0.118] |
| September | 1.064 [1.042-1.084] | 0.094 [0.078-0.113] |
| October | 1.088 [1.067-1.108] | 0.145 [0.120-0.173] |
| November | 1.063 [1.043-1.081] | 0.063 [0.053-0.074] |
| December | 1.011 [0.987-1.035] | 0.011 [0.009-0.013] |

**Table S17A. Results of parameter estimations with constant mutation probability and constant testing probability (Switzerland).** Samples of posterior distributions of the effective reproduction number and the dispersion parameter summarized by mean and 95% credible interval.

| Month | Mutation probability | Testing probability | Detection probability |
|---|---|---|---|
| January | 0.281 | 0.576 | 0.063 |
| February | 0.281 | 0.576 | 0.076 |
| March | 0.281 | 0.576 | 0.067 |
| April | 0.281 | 0.576 | 0.074 |
| May | 0.281 | 0.576 | 0.098 |
| June | 0.281 | 0.576 | 0.124 |
| July | 0.281 | 0.576 | 0.195 |
| August | 0.281 | 0.576 | 0.111 |
| September | 0.281 | 0.350 | 0.055 |
| October | 0.281 | 0.350 | 0.089 |
| November | 0.281 | 0.350 | 0.034 |
| December | 0.281 | 0.350 | 0.012 |

**Table S17B. Results of parameter estimations with constant mutation probability and constant testing probability (Switzerland).** Constant input values of the mutation probability, the testing probability and the detection probability.

##### 7.3.2 Denmark

| Month | Effective reproduction number | Dispersion parameter |
|---|---|---|
| January | 1.242 [1.227-1.258] | 0.378 [0.324-0.443] |
| February | 1.284 [1.267-1.302] | 0.559 [0.460-0.691] |
| March | 1.233 [1.214-1.251] | 0.492 [0.424-0.570] |
| April | 1.264 [1.248-1.280] | 0.499 [0.425-0.575] |
| May | 1.213 [1.195-1.230] | 0.314 [0.280-0.355] |
| June | 1.309 [1.290-1.328] | 0.319 [0.251-0.404] |
| July | 1.099 [1.077-1.120] | 0.245 [0.222-0.271] |
| August | 1.071 [1.055-1.087] | 0.385 [0.348-0.423] |
| September | 1.191 [1.174-1.209] | 0.332 [0.285-0.389] |
| October | 1.238 [1.225-1.250] | 0.319 [0.283-0.358] |
| November | 1.193 [1.183-1.202] | 0.166 [0.154-0.181] |
| December | 1.247 [1.238-1.256] | 0.072 [0.064-0.080] |

**Table S18A. Results of parameter estimations with constant mutation probability and constant testing probability (Denmark).** Samples of posterior distributions of the effective reproduction number and the dispersion parameter summarized by mean and 95% credible interval.

| Month | Mutation probability | Testing probability | Detection probability |
|---|---|---|---|
| January | 0.281 | 0.576 | 0.266 |
| February | 0.281 | 0.576 | 0.415 |
| March | 0.281 | 0.576 | 0.487 |
| April | 0.281 | 0.576 | 0.412 |
| May | 0.281 | 0.576 | 0.449 |
| June | 0.281 | 0.576 | 0.251 |
| July | 0.281 | 0.576 | 0.479 |
| August | 0.281 | 0.576 | 0.473 |
| September | 0.281 | 0.350 | 0.301 |
| October | 0.281 | 0.350 | 0.274 |
| November | 0.281 | 0.350 | 0.163 |
| December | 0.281 | 0.350 | 0.041 |

**Table S18B. Results of parameter estimations with constant mutation probability and constant testing probability (Denmark).** Constant input values of the mutation probability, the testing probability and the detection probability.

##### 7.3.3 Germany

| Month | Effective reproduction number | Dispersion parameter |
|---|---|---|
| January | 1.095 [1.064-1.122] | 0.014 [0.011-0.018] |
| February | 1.111 [1.095-1.125] | 0.086 [0.074-0.099] |
| March | 1.150 [1.139-1.160] | 0.145 [0.129-0.164] |
| April | 1.149 [1.139-1.159] | 0.142 [0.126-0.161] |
| May | 1.135 [1.124-1.147] | 0.158 [0.139-0.179] |
| June | 1.111 [1.091-1.129] | 0.222 [0.180-0.272] |
| July | 1.056 [1.034-1.076] | 0.190 [0.159-0.227] |
| August | 1.001 [0.989-1.014] | 0.148 [0.133-0.163] |
| September | 1.064 [1.054-1.075] | 0.130 [0.117-0.145] |
| October | 1.109 [1.098-1.122] | 0.074 [0.066-0.085] |
| November | 1.069 [1.056-1.081] | 0.038 [0.034-0.043] |
| December | 0.972 [0.962-0.984] | 0.014 [0.013-0.015] |

**Table S19A. Results of parameter estimations with constant mutation probability and constant testing probability (Germany).** Samples of posterior distributions of the effective reproduction number and the dispersion parameter summarized by mean and 95% credible interval.

| Month | Mutation probability | Testing probability | Detection probability |
|---|---|---|---|
| January | 0.281 | 0.576 | 0.008 |
| February | 0.281 | 0.576 | 0.047 |
| March | 0.281 | 0.576 | 0.053 |
| April | 0.281 | 0.576 | 0.042 |
| May | 0.281 | 0.576 | 0.052 |
| June | 0.281 | 0.576 | 0.097 |
| July | 0.281 | 0.576 | 0.120 |
| August | 0.281 | 0.576 | 0.095 |
| September | 0.281 | 0.350 | 0.050 |
| October | 0.281 | 0.350 | 0.029 |
| November | 0.281 | 0.350 | 0.013 |
| December | 0.281 | 0.350 | 0.014 |

**Table S19B. Results of parameter estimations with constant mutation probability and constant testing probability (Germany).** Constant input values of the mutation probability, the testing probability and the detection probability.

#### 7.4 Prior distribution for yearly mutation rate and prior distribution for testing probability

We use a scaled beta distribution with shape parameters $\alpha = 1$ and $\beta = 3$ defined on the interval $[0.05, 1]$ as prior distribution for the yearly mutation rate $M$ and the same prior distribution for the testing probability as in the original model. The following figures and tables present the results.

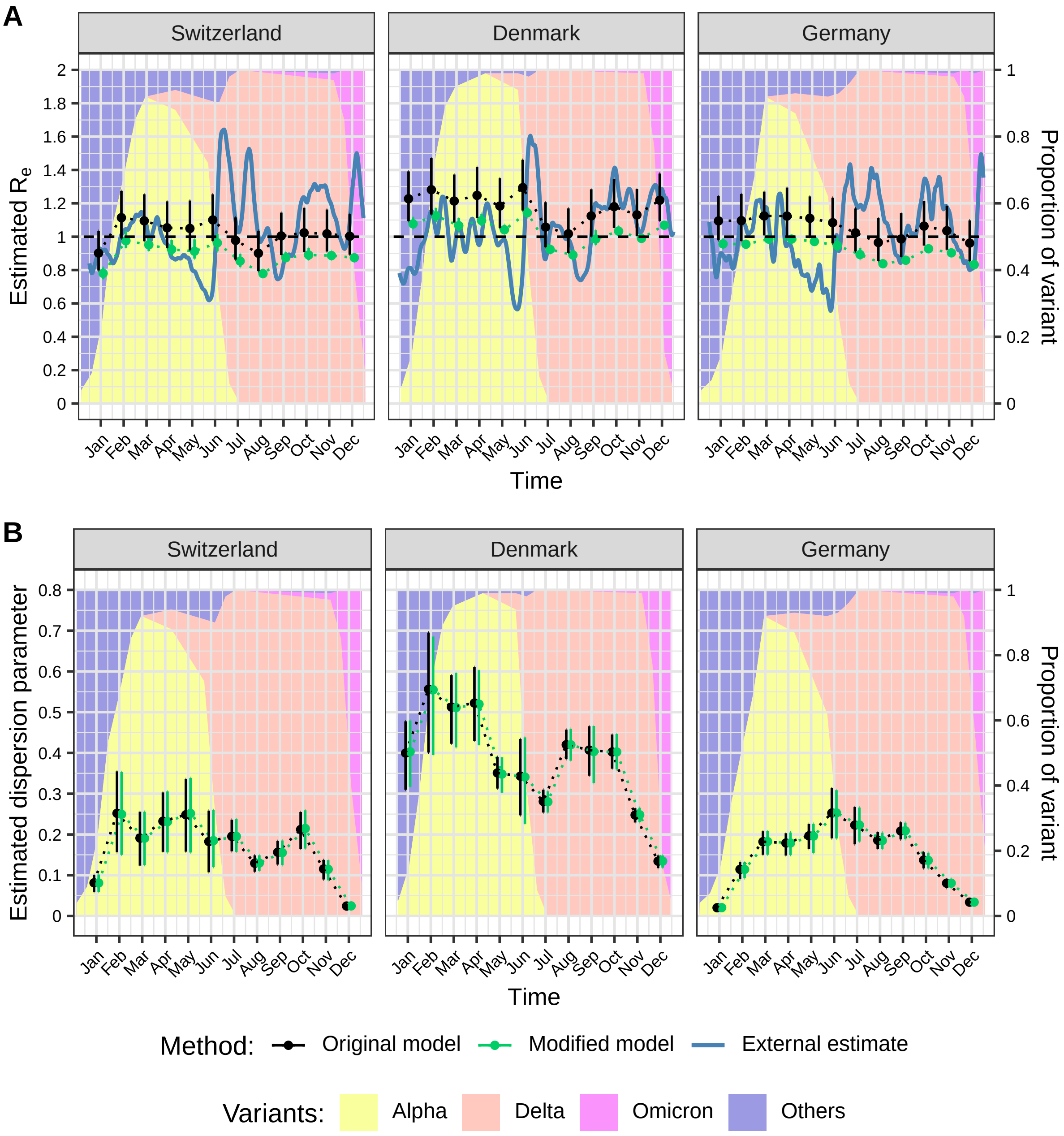

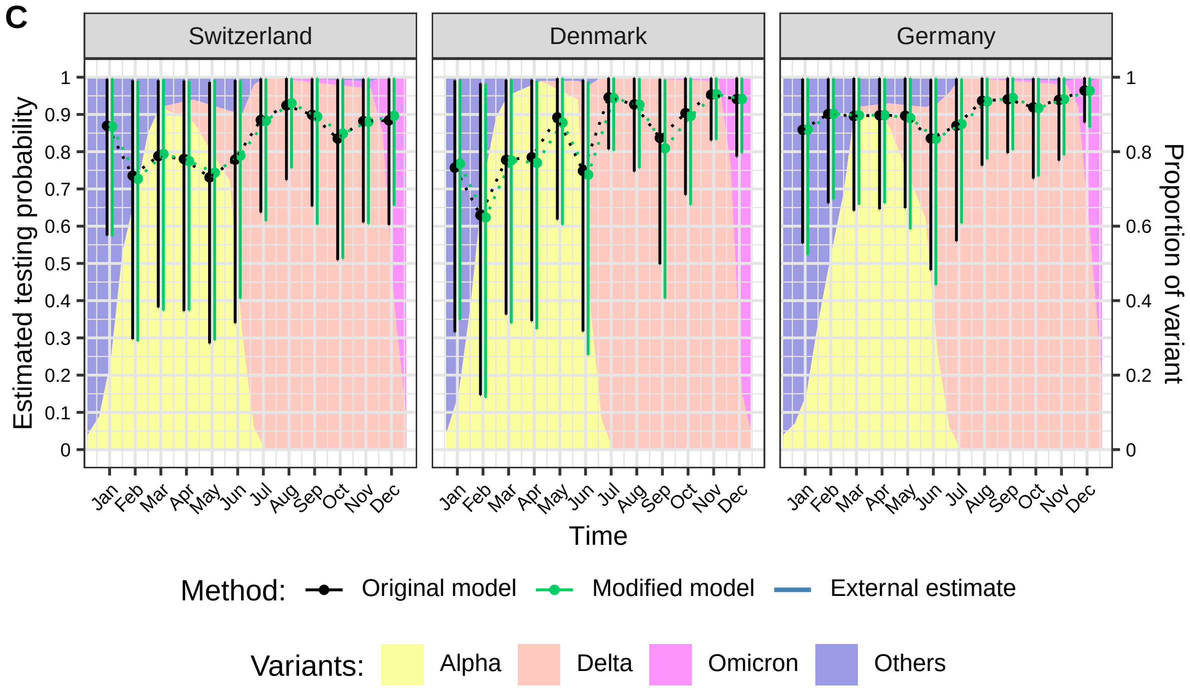

**Figure S8. Comparison of the estimated effective reproduction numbers, dispersion parameters and testing probabilities by country and month.** A: Effective reproduction number $R_e$. B: Dispersion parameter $k$. C: Testing probability $\tau_{test}$. The estimates are based on monthly time windows of identical sequence clusters. For each month the estimated means and 95% credible intervals of the posterior distribution obtained by two different models are shown. $R_e$ values are compared to external estimates based on laboratory-confirmed cases (in blue, from github.com/covid-19-Re [12]).

##### 7.4.1 Switzerland

| Month | Effective reproduction number | Dispersion parameter | Testing probability |
|---|---|---|---|
| January | 0.780 [0.748-0.816] | 0.081 [0.059-0.102] | 0.867 [0.574-0.997] |
| February | 0.976 [0.932-1.036] | 0.250 [0.150-0.353] | 0.727 [0.291-0.991] |
| March | 0.951 [0.914-1.000] | 0.191 [0.125-0.256] | 0.794 [0.374-0.994] |
| April | 0.924 [0.886-0.981] | 0.231 [0.156-0.306] | 0.775 [0.374-0.990] |
| May | 0.913 [0.869-0.984] | 0.252 [0.156-0.339] | 0.744 [0.293-0.993] |
| June | 0.963 [0.913-1.017] | 0.185 [0.120-0.260] | 0.790 [0.406-0.991] |
| July | 0.854 [0.817-0.897] | 0.195 [0.157-0.238] | 0.883 [0.614-0.996] |
| August | 0.778 [0.755-0.805] | 0.130 [0.111-0.149] | 0.930 [0.756-0.997] |
| September | 0.874 [0.848-0.908] | 0.155 [0.125-0.185] | 0.894 [0.604-0.998] |
| October | 0.890 [0.860-0.933] | 0.215 [0.165-0.259] | 0.848 [0.512-0.996] |
| November | 0.887 [0.863-0.914] | 0.115 [0.087-0.137] | 0.880 [0.606-0.997] |
| December | 0.874 [0.851-0.896] | 0.025 [0.018-0.030] | 0.896 [0.655-0.998] |

**Table S20A. Results of parameter estimations (Switzerland).** Samples of posterior distributions of the effective reproduction number, the dispersion parameter and the testing probability summarized by mean and 95% credible interval.

| Month | Yearly mutation rate | Mutation probability | Detection probability |
|---|---|---|---|
| January | 14.002 [13.016-14.982] | 0.181 [0.169-0.192] | 0.096 [0.063-0.110] |
| February | 14.009 [13.063-14.976] | 0.181 [0.170-0.192] | 0.095 [0.038-0.130] |
| March | 14.004 [12.956-14.958] | 0.181 [0.168-0.192] | 0.092 [0.044-0.116] |
| April | 13.996 [13.013-14.940] | 0.181 [0.169-0.192] | 0.100 [0.048-0.127] |
| May | 13.986 [13.091-14.920] | 0.181 [0.170-0.191] | 0.127 [0.050-0.169] |
| June | 13.996 [12.967-14.998] | 0.181 [0.169-0.192] | 0.170 [0.087-0.213] |
| July | 14.011 [13.107-14.967] | 0.181 [0.170-0.192] | 0.299 [0.208-0.337] |
| August | 14.016 [13.070-14.962] | 0.181 [0.170-0.192] | 0.179 [0.146-0.192] |
| September | 13.993 [12.998-14.973] | 0.181 [0.169-0.192] | 0.140 [0.095-0.156] |
| October | 13.983 [13.042-14.973] | 0.180 [0.169-0.192] | 0.216 [0.131-0.254] |
| November | 13.988 [13.013-14.918] | 0.181 [0.169-0.191] | 0.086 [0.059-0.097] |
| December | 13.994 [12.966-14.982] | 0.181 [0.169-0.192] | 0.030 [0.022-0.033] |

**Table S20B. Results of parameter estimations (Switzerland).** Samples of posterior distributions of the yearly mutation rate, the mutation probability and the detection probability summarized by mean and 95% credible interval.

##### 7.4.2 Denmark

| Month | Effective reproduction number | Dispersion parameter | Testing probability |
|---|---|---|---|
| January | 1.077 [1.048-1.116] | 0.403 [0.317-0.480] | 0.768 [0.349-0.994] |
| February | 1.126 [1.088-1.173] | 0.555 [0.395-0.685] | 0.624 [0.140-0.982] |
| March | 1.066 [1.033-1.111] | 0.511 [0.414-0.596] | 0.777 [0.339-0.994] |
| April | 1.096 [1.066-1.138] | 0.520 [0.420-0.603] | 0.770 [0.324-0.990] |
| May | 1.041 [1.015-1.072] | 0.348 [0.303-0.389] | 0.878 [0.604-0.998] |
| June | 1.143 [1.116-1.175] | 0.341 [0.226-0.438] | 0.738 [0.254-0.988] |
| July | 0.924 [0.898-0.951] | 0.280 [0.254-0.306] | 0.943 [0.803-0.998] |
| August | 0.890 [0.867-0.918] | 0.420 [0.381-0.459] | 0.926 [0.756-0.997] |
| September | 0.986 [0.950-1.040] | 0.403 [0.326-0.466] | 0.809 [0.406-0.994] |
| October | 1.033 [1.009-1.062] | 0.403 [0.360-0.446] | 0.896 [0.657-0.998] |
| November | 0.990 [0.972-1.009] | 0.248 [0.230-0.266] | 0.954 [0.832-0.999] |
| December | 1.069 [1.052-1.086] | 0.135 [0.118-0.149] | 0.941 [0.796-0.998] |

**Table S21A. Results of parameter estimations (Denmark).** Samples of posterior distributions of the effective reproduction number, the dispersion parameter and the testing probability summarized by mean and 95% credible interval.

| Month | Yearly mutation rate | Mutation probability | Detection probability |
|---|---|---|---|
| January | 14.030 [13.071-14.993] | 0.181 [0.170-0.192] | 0.354 [0.161-0.458] |
| February | 14.008 [13.049-14.949] | 0.181 [0.170-0.192] | 0.449 [0.101-0.708] |
| March | 14.016 [12.964-15.019] | 0.181 [0.169-0.193] | 0.656 [0.286-0.840] |
| April | 14.024 [12.999-15.031] | 0.181 [0.169-0.193] | 0.552 [0.232-0.709] |
| May | 14.022 [13.101-14.962] | 0.181 [0.170-0.192] | 0.684 [0.470-0.777] |
| June | 13.997 [12.987-15.011] | 0.181 [0.169-0.192] | 0.322 [0.111-0.431] |
| July | 14.030 [13.077-14.957] | 0.181 [0.170-0.192] | 0.784 [0.667-0.830] |
| August | 14.008 [13.004-15.014] | 0.181 [0.169-0.192] | 0.761 [0.621-0.820] |
| September | 13.993 [12.920-14.935] | 0.181 [0.168-0.192] | 0.695 [0.349-0.854] |
| October | 14.003 [12.983-14.976] | 0.181 [0.169-0.192] | 0.703 [0.515-0.782] |
| November | 13.988 [13.038-14.976] | 0.181 [0.169-0.192] | 0.444 [0.387-0.464] |
| December | 13.992 [13.050-14.920] | 0.181 [0.170-0.191] | 0.110 [0.093-0.117] |

**Table S21B. Results of parameter estimations (Denmark).** Samples of posterior distributions of the yearly mutation rate, the mutation probability and the detection probability summarized by mean and 95% credible interval.

##### 7.4.3 Germany

| Month | Effective reproduction number | Dispersion parameter | Testing probability |
|---|---|---|---|
| January | 0.957 [0.929-0.986] | 0.020 [0.013-0.028] | 0.860 [0.523-0.997] |
| February | 0.955 [0.935-0.975] | 0.114 [0.092-0.132] | 0.902 [0.672-0.997] |
| March | 0.985 [0.966-1.009] | 0.183 [0.150-0.208] | 0.898 [0.658-0.998] |
| April | 0.985 [0.967-1.006] | 0.179 [0.149-0.205] | 0.898 [0.662-0.997] |
| May | 0.971 [0.950-0.997] | 0.196 [0.154-0.226] | 0.890 [0.592-0.998] |
| June | 0.948 [0.916-0.996] | 0.253 [0.190-0.309] | 0.835 [0.442-0.997] |
| July | 0.893 [0.864-0.933] | 0.223 [0.183-0.266] | 0.874 [0.608-0.996] |
| August | 0.838 [0.820-0.859] | 0.185 [0.165-0.204] | 0.935 [0.779-0.998] |
| September | 0.859 [0.843-0.879] | 0.209 [0.188-0.229] | 0.944 [0.805-0.999] |
| October | 0.927 [0.909-0.948] | 0.136 [0.116-0.155] | 0.917 [0.734-0.997] |
| November | 0.903 [0.886-0.921] | 0.080 [0.069-0.091] | 0.941 [0.791-0.998] |
| December | 0.832 [0.818-0.847] | 0.034 [0.030-0.037] | 0.963 [0.865-0.998] |

**Table S22A. Results of parameter estimations (Germany).** Samples of posterior distributions of the effective reproduction number, the dispersion parameter and the testing probability summarized by mean and 95% credible interval.

| Month | Yearly mutation rate | Mutation probability | Detection probability |
|---|---|---|---|
| January | 14.041 [13.101-15.026] | 0.181 [0.170-0.193] | 0.012 [0.007-0.014] |
| February | 13.990 [12.999-14.888] | 0.181 [0.169-0.191] | 0.073 [0.055-0.081] |
| March | 14.004 [13.050-15.007] | 0.181 [0.170-0.192] | 0.083 [0.061-0.092] |
| April | 14.032 [12.965-14.993] | 0.181 [0.169-0.192] | 0.066 [0.048-0.073] |
| May | 13.986 [13.031-14.967] | 0.181 [0.169-0.192] | 0.081 [0.054-0.091] |
| June | 14.006 [13.043-15.017] | 0.181 [0.169-0.192] | 0.141 [0.075-0.168] |
| July | 14.012 [12.998-14.996] | 0.181 [0.169-0.192] | 0.183 [0.127-0.208] |
| August | 13.992 [13.098-15.029] | 0.181 [0.170-0.193] | 0.154 [0.129-0.165] |
| September | 13.996 [13.093-15.012] | 0.181 [0.170-0.192] | 0.135 [0.115-0.143] |
| October | 14.017 [13.020-14.973] | 0.181 [0.169-0.192] | 0.076 [0.061-0.083] |
| November | 13.951 [12.994-15.005] | 0.180 [0.169-0.192] | 0.035 [0.030-0.038] |
| December | 14.009 [13.003-15.045] | 0.181 [0.169-0.193] | 0.038 [0.034-0.039] |

**Table S22B. Results of parameter estimations (Germany).** Samples of posterior distributions of the yearly mutation rate, the mutation probability and the detection probability summarized by mean and 95% credible interval.

#### 7.5 Prior distribution for yearly mutation rate and constant testing probability

We use a scaled beta distribution with shape parameters $\alpha = 1$ and $\beta = 3$ defined on the interval $[0.05, 1]$ as prior distribution for the yearly mutation rate $M$. Instead of a prior distribution for the testing probability, constant values are used. The following figures and tables present the results.

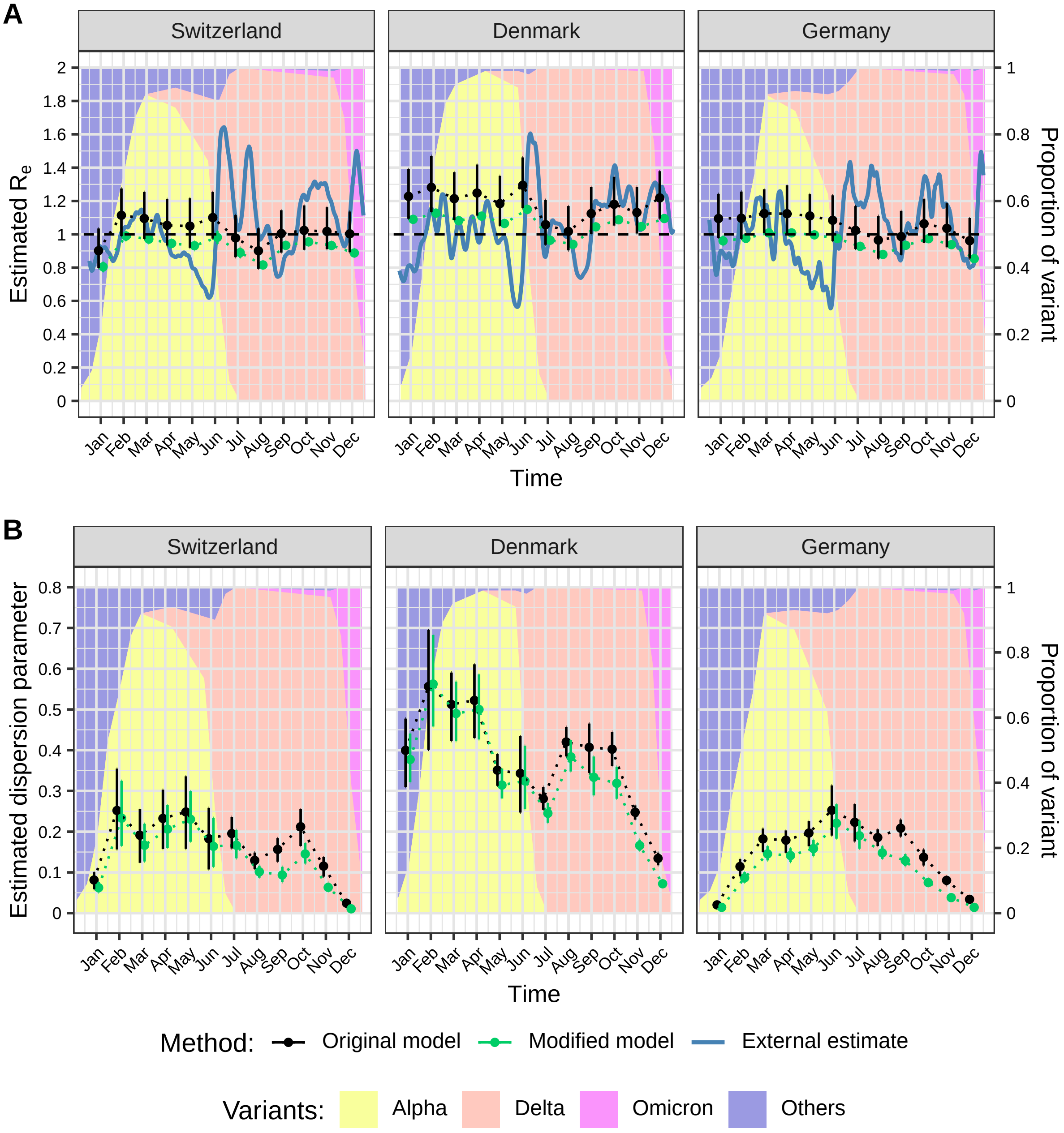

**Figure S9. Comparison of the estimated effective reproduction numbers and dispersion parameters by country and month.** A: Effective reproduction number $R_e$. B: Dispersion parameter $k$. The estimates are based on monthly time windows of identical sequence clusters. For each month the estimated means and 95% credible intervals of the posterior distribution obtained by two different models are shown. $R_e$ values are compared to external estimates based on laboratory-confirmed cases (in blue, from github.com/covid-19-Re [12]).

##### 7.5.1 Switzerland

| Month | Effective reproduction number | Dispersion parameter | Testing probability |
|---|---|---|---|
| January | 0.805 [0.775-0.834] | 0.062 [0.051-0.076] | 0.576 |
| February | 0.990 [0.960-1.019] | 0.233 [0.165-0.325] | 0.576 |
| March | 0.971 [0.944-0.997] | 0.166 [0.127-0.219] | 0.576 |
| April | 0.945 [0.920-0.969] | 0.206 [0.161-0.265] | 0.576 |
| May | 0.932 [0.905-0.957] | 0.230 [0.175-0.299] | 0.576 |
| June | 0.982 [0.942-1.021] | 0.164 [0.114-0.234] | 0.576 |
| July | 0.890 [0.859-0.920] | 0.167 [0.135-0.204] | 0.576 |
| August | 0.816 [0.794-0.838] | 0.101 [0.087-0.117] | 0.576 |
| September | 0.934 [0.911-0.956] | 0.094 [0.076-0.114] | 0.350 |
| October | 0.955 [0.932-0.976] | 0.145 [0.122-0.172] | 0.350 |
| November | 0.933 [0.911-0.955] | 0.063 [0.053-0.075] | 0.350 |
| December | 0.888 [0.866-0.912] | 0.011 [0.009-0.013] | 0.350 |

**Table S23A. Results of parameter estimations with constant testing probability (Switzerland).** Samples of posterior distributions of the effective reproduction number and the dispersion parameter summarized by mean and 95% credible interval.

| Month | Yearly mutation rate | Mutation probability | Detection probability |
|---|---|---|---|
| January | 14.006 [13.036-15.000] | 0.181 [0.169-0.192] | 0.063 |
| February | 13.998 [13.041-15.006] | 0.181 [0.169-0.192] | 0.076 |
| March | 14.008 [13.000-14.928] | 0.181 [0.169-0.191] | 0.067 |
| April | 14.013 [13.062-14.939] | 0.181 [0.170-0.192] | 0.074 |
| May | 13.969 [13.045-14.925] | 0.180 [0.169-0.191] | 0.098 |
| June | 13.988 [13.037-14.919] | 0.181 [0.169-0.191] | 0.124 |
| July | 13.996 [13.053-14.984] | 0.181 [0.170-0.192] | 0.195 |
| August | 13.987 [13.035-15.005] | 0.181 [0.169-0.192] | 0.111 |
| September | 13.998 [13.089-14.965] | 0.181 [0.170-0.192] | 0.055 |
| October | 13.980 [13.001-14.958] | 0.180 [0.169-0.192] | 0.089 |
| November | 14.010 [12.997-15.035] | 0.181 [0.169-0.193] | 0.034 |
| December | 14.008 [13.078-14.973] | 0.181 [0.170-0.192] | 0.012 |

**Table S23B. Results of parameter estimations with constant testing probability (Switzerland).** Samples of posterior distributions of the yearly mutation rate and the mutation probability summarized by mean and 95% credible interval.

##### 7.5.2 Denmark

| Month | Effective reproduction number | Dispersion parameter | Testing probability |
|---|---|---|---|
| January | 1.090 [1.070-1.110] | 0.377 [0.321-0.441] | 0.576 |
| February | 1.127 [1.105-1.149] | 0.562 [0.458-0.683] | 0.576 |
| March | 1.082 [1.058-1.105] | 0.490 [0.422-0.568] | 0.576 |
| April | 1.109 [1.088-1.130] | 0.500 [0.426-0.586] | 0.576 |
| May | 1.065 [1.043-1.086] | 0.314 [0.281-0.355] | 0.576 |
| June | 1.149 [1.126-1.173] | 0.324 [0.255-0.411] | 0.576 |
| July | 0.964 [0.941-0.988] | 0.245 [0.222-0.271] | 0.576 |
| August | 0.940 [0.921-0.958] | 0.384 [0.348-0.425] | 0.576 |
| September | 1.045 [1.024-1.067] | 0.333 [0.289-0.385] | 0.350 |
| October | 1.087 [1.069-1.107] | 0.319 [0.281-0.359] | 0.350 |
| November | 1.047 [1.029-1.064] | 0.166 [0.153-0.180] | 0.350 |
| December | 1.095 [1.077-1.111] | 0.072 [0.064-0.081] | 0.350 |

**Table S24A. Results of parameter estimations with constant testing probability (Denmark).** Samples of posterior distributions of the effective reproduction number and the dispersion parameter summarized by mean and 95% credible interval.

| Month | Yearly mutation rate | Mutation probability | Detection probability |
|---|---|---|---|
| January | 13.979 [13.050-14.925] | 0.180 [0.170-0.191] | 0.266 |
| February | 13.978 [12.993-14.963] | 0.180 [0.169-0.192] | 0.415 |
| March | 13.987 [12.933-14.946] | 0.181 [0.168-0.192] | 0.487 |
| April | 14.005 [13.058-14.970] | 0.181 [0.170-0.192] | 0.412 |
| May | 14.007 [13.042-14.988] | 0.181 [0.169-0.192] | 0.449 |
| June | 14.006 [13.023-15.025] | 0.181 [0.169-0.193] | 0.251 |
| July | 14.004 [12.982-14.969] | 0.181 [0.169-0.192] | 0.479 |
| August | 13.983 [12.996-14.964] | 0.180 [0.169-0.192] | 0.473 |
| September | 13.979 [13.021-14.918] | 0.180 [0.169-0.191] | 0.301 |
| October | 14.043 [13.099-15.041] | 0.181 [0.170-0.193] | 0.274 |
| November | 13.997 [12.996-15.026] | 0.181 [0.169-0.193] | 0.163 |
| December | 14.013 [13.053-14.927] | 0.181 [0.170-0.191] | 0.041 |

**Table S24B. Results of parameter estimations with constant testing probability (Denmark).** Samples of posterior distributions of the yearly mutation rate and the mutation probability summarized by mean and 95% credible interval.

##### 7.5.3 Germany

| Month | Effective reproduction number | Dispersion parameter | Testing probability |
|---|---|---|---|
| January | 0.961 [0.932-0.991] | 0.014 [0.011-0.018] | 0.576 |
| February | 0.976 [0.957-0.994] | 0.087 [0.075-0.101] | 0.576 |
| March | 1.009 [0.991-1.026] | 0.145 [0.129-0.164] | 0.576 |
| April | 1.008 [0.992-1.025] | 0.141 [0.125-0.159] | 0.576 |
| May | 0.997 [0.979-1.013] | 0.159 [0.140-0.180] | 0.576 |
| June | 0.975 [0.953-0.996] | 0.221 [0.183-0.267] | 0.576 |
| July | 0.928 [0.905-0.951] | 0.190 [0.158-0.227] | 0.576 |
| August | 0.879 [0.863-0.895] | 0.148 [0.133-0.164] | 0.576 |
| September | 0.934 [0.918-0.951] | 0.129 [0.115-0.145] | 0.350 |
| October | 0.974 [0.956-0.992] | 0.075 [0.066-0.085] | 0.350 |
| November | 0.938 [0.920-0.956] | 0.038 [0.034-0.043] | 0.350 |
| December | 0.854 [0.838-0.869] | 0.014 [0.013-0.015] | 0.350 |

**Table S25A. Results of parameter estimations with constant testing probability (Germany).** Samples of posterior distributions of the effective reproduction number and the dispersion parameter summarized by mean and 95% credible interval.

| Month | Yearly mutation rate | Mutation probability | Detection probability |
|---|---|---|---|
| January | 14.012 [13.074-15.007] | 0.181 [0.170-0.192] | 0.008 |
| February | 14.002 [12.981-14.943] | 0.181 [0.169-0.192] | 0.047 |
| March | 13.989 [13.030-14.994] | 0.181 [0.169-0.192] | 0.053 |
| April | 14.011 [13.022-15.013] | 0.181 [0.169-0.192] | 0.042 |
| May | 14.003 [12.990-14.956] | 0.181 [0.169-0.192] | 0.052 |
| June | 13.999 [13.057-14.982] | 0.181 [0.170-0.192] | 0.097 |
| July | 14.032 [13.083-15.020] | 0.181 [0.170-0.193] | 0.120 |
| August | 14.032 [13.061-14.963] | 0.181 [0.170-0.192] | 0.095 |
| September | 14.010 [13.021-15.013] | 0.181 [0.169-0.192] | 0.050 |
| October | 13.965 [12.993-14.988] | 0.180 [0.169-0.192] | 0.029 |
| November | 14.001 [13.048-15.001] | 0.181 [0.170-0.192] | 0.013 |
| December | 14.005 [13.033-14.973] | 0.181 [0.169-0.192] | 0.014 |

**Table S25B. Results of parameter estimations with constant testing probability (Germany).** Samples of posterior distributions of the yearly mutation rate and the mutation probability summarized by mean and 95% credible interval.

### 8 Comparison with literature

In order to compare our model with the work of Tran-Kiem and Bedford [13], we apply our model to the same data set from New Zealand as they did and also use the same parameters. In order to achieve the highest possible comparability, we use constant values for both the mutation probability and the testing probability. We retrieved the cluster data as well as the values for mutation probability, testing probability and sequencing probability from the GitHub repository size-genetic-clusters [14]. We have obtained very similar results and present them in the following figure in the same way as did Tran-Kiem and Bedford in figure S9 of the supplementary information of [13].

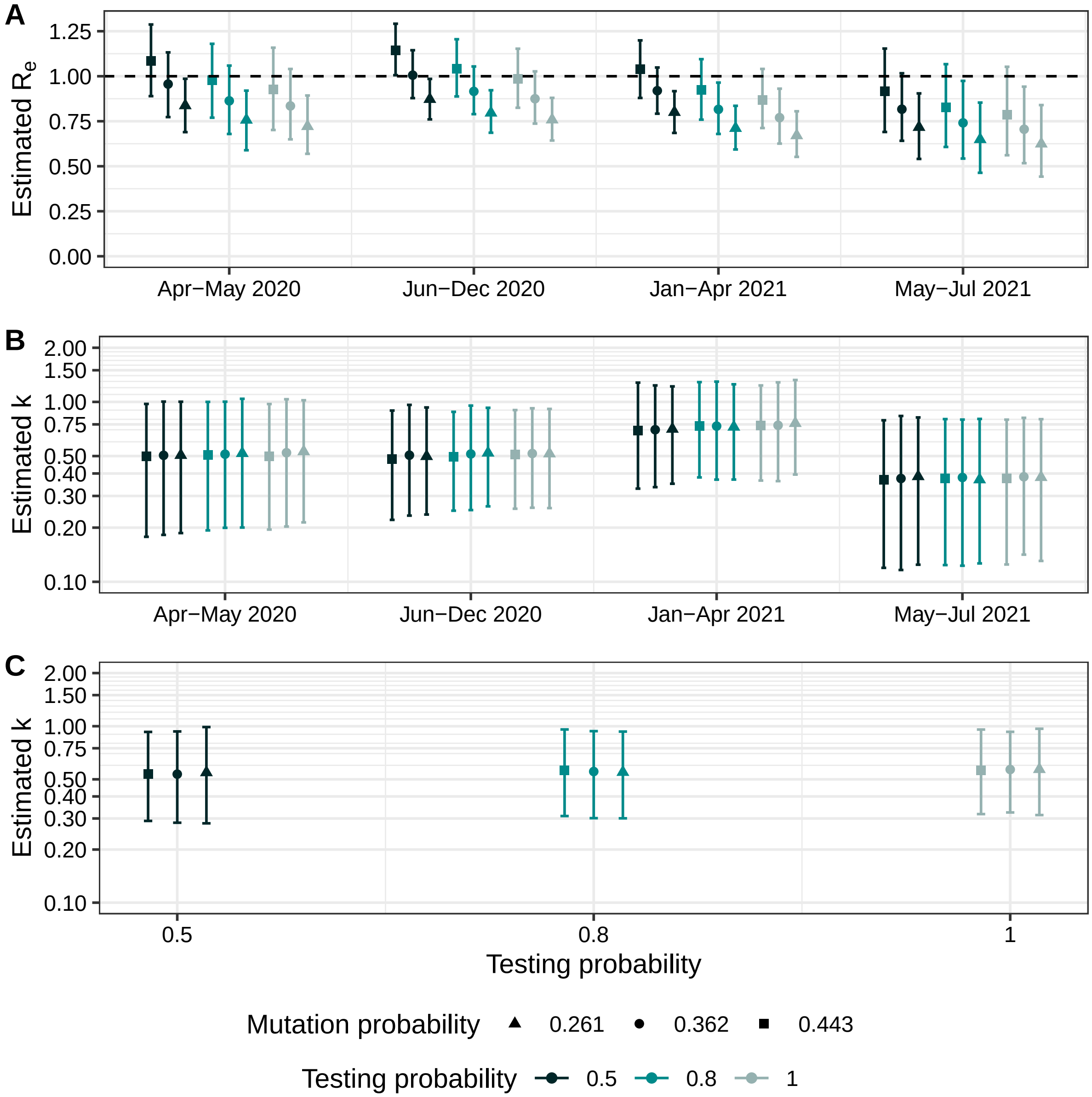

**Figure S10. Comparison of the estimated effective reproduction numbers and dispersion parameters in New Zealand for different timne periods.** A: Effective reproduction number $R_e$ by period. B: Dispersion parameter $k$ by period. C: Dispersion parameter $k$ over whole time period. For each period and combination of mutation probability and testing probability the estimated means and 95% credible intervals of the posterior distribution are shown.

| Index | Period | Effective reproduction number | Dispersion parameter |
|---|---|---|---|
| 1 | Apr-May 2020 | 0.837 [0.689-0.986] | 0.506 [0.187-1.003] |
| 2 | Jun-Dec 2020 | 0.872 [0.760-0.985] | 0.498 [0.237-0.932] |
| 3 | Jan-Apr 2021 | 0.800 [0.685-0.917] | 0.707 [0.351-1.220] |
| 4 | May-Jul 2021 | 0.717 [0.541-0.905] | 0.385 [0.124-0.820] |
| 5 | Apr-May 2020 | 0.956 [0.773-1.133] | 0.505 [0.182-1.004] |
| 6 | Jun-Dec 2020 | 1.006 [0.878-1.144] | 0.505 [0.233-0.963] |
| 7 | Jan-Apr 2021 | 0.920 [0.792-1.048] | 0.700 [0.336-1.235] |
| 8 | May-Jul 2021 | 0.817 [0.641-1.016] | 0.375 [0.116-0.835] |
| 9 | Apr-May 2020 | 1.086 [0.889-1.287] | 0.499 [0.178-0.974] |
| 10 | Jun-Dec 2020 | 1.144 [1.006-1.292] | 0.481 [0.221-0.895] |
| 11 | Jan-Apr 2021 | 1.039 [0.879-1.199] | 0.692 [0.329-1.280] |
| 12 | May-Jul 2021 | 0.916 [0.690-1.154] | 0.368 [0.120-0.790] |
| 13 | Apr-May 2020 | 0.757 [0.589-0.920] | 0.518 [0.200-1.040] |
| 14 | Jun-Dec 2020 | 0.796 [0.686-0.922] | 0.521 [0.263-0.927] |
| 15 | Jan-Apr 2021 | 0.711 [0.593-0.836] | 0.726 [0.370-1.253] |
| 16 | May-Jul 2021 | 0.649 [0.464-0.854] | 0.370 [0.127-0.804] |
| 17 | Apr-May 2020 | 0.863 [0.679-1.059] | 0.512 [0.200-1.003] |
| 18 | Jun-Dec 2020 | 0.916 [0.789-1.054] | 0.514 [0.250-0.954] |
| 19 | Jan-Apr 2021 | 0.816 [0.679-0.965] | 0.733 [0.370-1.296] |
| 20 | May-Jul 2021 | 0.741 [0.543-0.974] | 0.380 [0.123-0.797] |
| 21 | Apr-May 2020 | 0.978 [0.770-1.180] | 0.506 [0.193-1.000] |
| 22 | Jun-Dec 2020 | 1.041 [0.888-1.205] | 0.495 [0.248-0.881] |
| 23 | Jan-Apr 2021 | 0.923 [0.759-1.095] | 0.733 [0.380-1.288] |
| 24 | May-Jul 2021 | 0.827 [0.607-1.067] | 0.375 [0.124-0.802] |

**Table S26A. Results of parameter estimations with constant mutation probability and constant testing probability (New Zealand).** Samples of posterior distributions of the effective reproduction number and the dispersion parameter summarized by mean and 95% credible interval.

| Index | Period | Effective reproduction number | Dispersion parameter |
|---|---|---|---|
| 25 | Apr-May 2020 | 0.722 [0.569-0.893] | 0.530 [0.214-1.021] |
| 26 | Jun-Dec 2020 | 0.759 [0.643-0.880] | 0.516 [0.257-0.914] |
| 27 | Jan-Apr 2021 | 0.671 [0.552-0.805] | 0.761 [0.395-1.324] |
| 28 | May-Jul 2021 | 0.625 [0.442-0.840] | 0.381 [0.131-0.802] |
| 29 | Apr-May 2020 | 0.835 [0.649-1.040] | 0.522 [0.203-1.035] |
| 30 | Jun-Dec 2020 | 0.875 [0.737-1.027] | 0.516 [0.258-0.922] |
| 31 | Jan-Apr 2021 | 0.770 [0.626-0.931] | 0.741 [0.363-1.285] |
| 32 | May-Jul 2021 | 0.705 [0.517-0.942] | 0.383 [0.142-0.816] |
| 33 | Apr-May 2020 | 0.926 [0.701-1.158] | 0.498 [0.195-0.973] |
| 34 | Jun-Dec 2020 | 0.984 [0.825-1.153] | 0.509 [0.255-0.901] |
| 35 | Jan-Apr 2021 | 0.868 [0.712-1.041] | 0.738 [0.365-1.235] |
| 36 | May-Jul 2021 | 0.787 [0.561-1.052] | 0.375 [0.125-0.797] |

**Table S26A. Results of parameter estimations with constant mutation probability and constant testing probability (New Zealand).** Samples of posterior distributions of the effective reproduction number and the dispersion parameter summarized by mean and 95% credible interval.

| Index | Period | Mutation probability | Testing probability | Detection probability |
|---|---|---|---|---|
| 1 | Apr-May 2020 | 0.261 | 0.500 | 0.126 |
| 2 | Jun-Dec 2020 | 0.261 | 0.500 | 0.254 |
| 3 | Jan-Apr 2021 | 0.261 | 0.500 | 0.229 |
| 4 | May-Jul 2021 | 0.261 | 0.500 | 0.240 |
| 5 | Apr-May 2020 | 0.362 | 0.500 | 0.126 |
| 6 | Jun-Dec 2020 | 0.362 | 0.500 | 0.254 |
| 7 | Jan-Apr 2021 | 0.362 | 0.500 | 0.229 |
| 8 | May-Jul 2021 | 0.362 | 0.500 | 0.240 |
| 9 | Apr-May 2020 | 0.443 | 0.500 | 0.126 |
| 10 | Jun-Dec 2020 | 0.443 | 0.500 | 0.254 |
| 11 | Jan-Apr 2021 | 0.443 | 0.500 | 0.229 |
| 12 | May-Jul 2021 | 0.443 | 0.500 | 0.240 |

**Table S26B. Results of parameter estimations with constant mutation probability and constant testing probability (New Zealand).** Constant input values of the mutation probability, the testing probability and the detection probability.

| Index | Period | Mutation probability | Testing probability | Detection probability |
|---|---|---|---|---|
| 13 | Apr-May 2020 | 0.261 | 0.800 | 0.202 |
| 14 | Jun-Dec 2020 | 0.261 | 0.800 | 0.407 |
| 15 | Jan-Apr 2021 | 0.261 | 0.800 | 0.367 |
| 16 | May-Jul 2021 | 0.261 | 0.800 | 0.384 |
| 17 | Apr-May 2020 | 0.362 | 0.800 | 0.202 |
| 18 | Jun-Dec 2020 | 0.362 | 0.800 | 0.407 |
| 19 | Jan-Apr 2021 | 0.362 | 0.800 | 0.367 |
| 20 | May-Jul 2021 | 0.362 | 0.800 | 0.384 |
| 21 | Apr-May 2020 | 0.443 | 0.800 | 0.202 |
| 22 | Jun-Dec 2020 | 0.443 | 0.800 | 0.407 |
| 23 | Jan-Apr 2021 | 0.443 | 0.800 | 0.367 |
| 24 | May-Jul 2021 | 0.443 | 0.800 | 0.384 |
| 25 | Apr-May 2020 | 0.261 | 1.000 | 0.252 |
| 26 | Jun-Dec 2020 | 0.261 | 1.000 | 0.508 |
| 27 | Jan-Apr 2021 | 0.261 | 1.000 | 0.459 |
| 28 | May-Jul 2021 | 0.261 | 1.000 | 0.480 |
| 29 | Apr-May 2020 | 0.362 | 1.000 | 0.252 |
| 30 | Jun-Dec 2020 | 0.362 | 1.000 | 0.508 |
| 31 | Jan-Apr 2021 | 0.362 | 1.000 | 0.459 |
| 32 | May-Jul 2021 | 0.362 | 1.000 | 0.480 |
| 33 | Apr-May 2020 | 0.443 | 1.000 | 0.252 |
| 34 | Jun-Dec 2020 | 0.443 | 1.000 | 0.508 |
| 35 | Jan-Apr 2021 | 0.443 | 1.000 | 0.459 |
| 36 | May-Jul 2021 | 0.443 | 1.000 | 0.480 |

**Table S26B. Results of parameter estimations with constant mutation probability and constant testing probability (New Zealand).** Constant input values of the mutation probability, the testing probability and the detection probability.

### 9 Posterior predictive check

We ran a posterior predictive check to assess whether our simulation of identical sequence clusters is compatible with the cluster data from Switzerland, Denmark and Germany. For each country and month we randomly chose 50 samples each from the posterior distributions of the effective reproduction number $R_e$, the dispersion parameter $k$, the mutation probability $\mu$ and the detection probability $\tau$ obtained with our main model (presented in the paper). We then simulated the same number of clusters observed in the respective country and month, running the simulation once for each of the 50 parameter combinations. We summarized the results of the simulations by the 95% prediction interval, calculated individually for each cluster size. The following figures present the outcome of the posterior predictive check.

#### 9.1 Switzerland

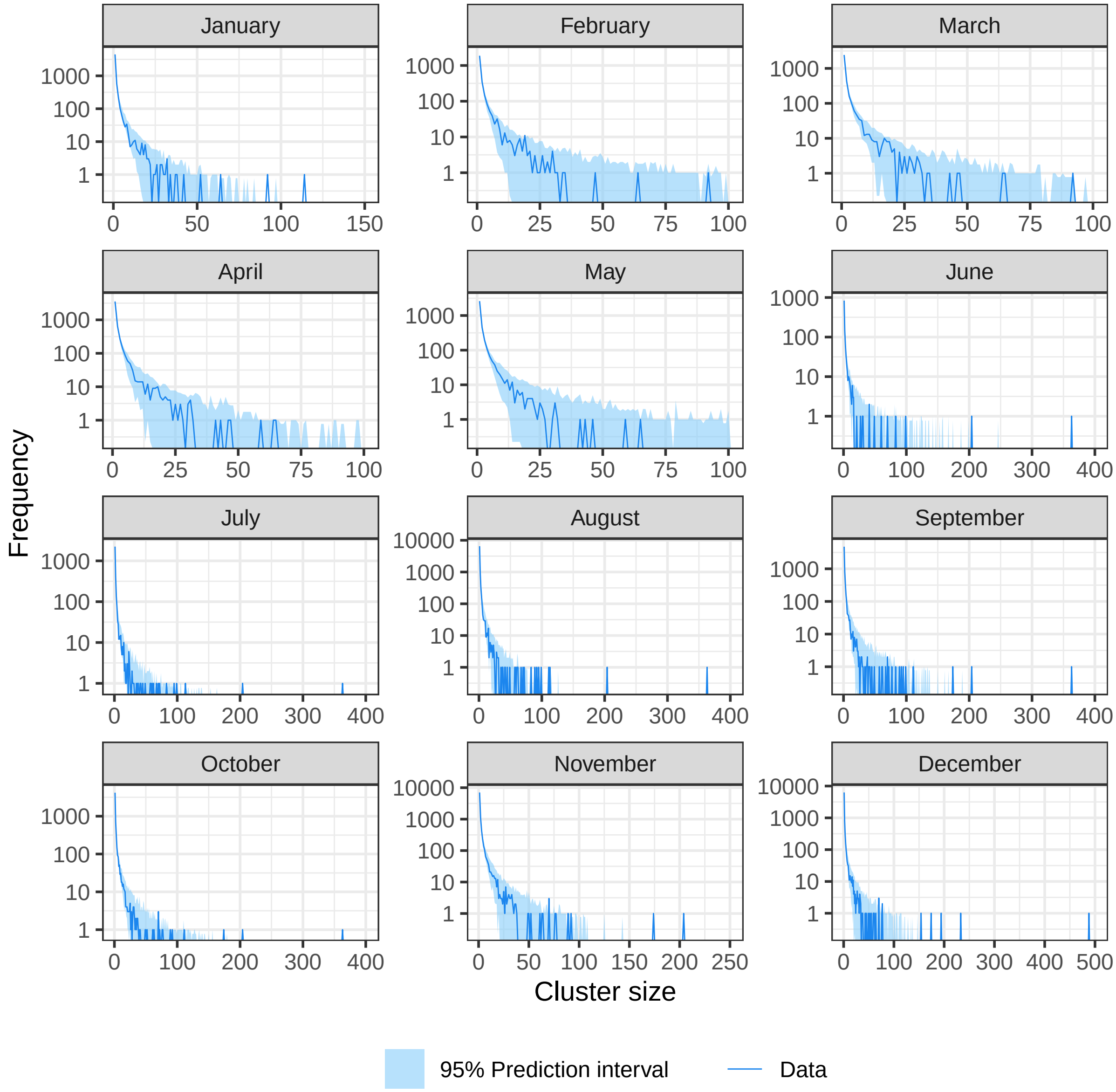

**Figure S11. Posterior predictive check of simulation of cluster size distribution (Switzerland).**

#### 9.2 Denmark

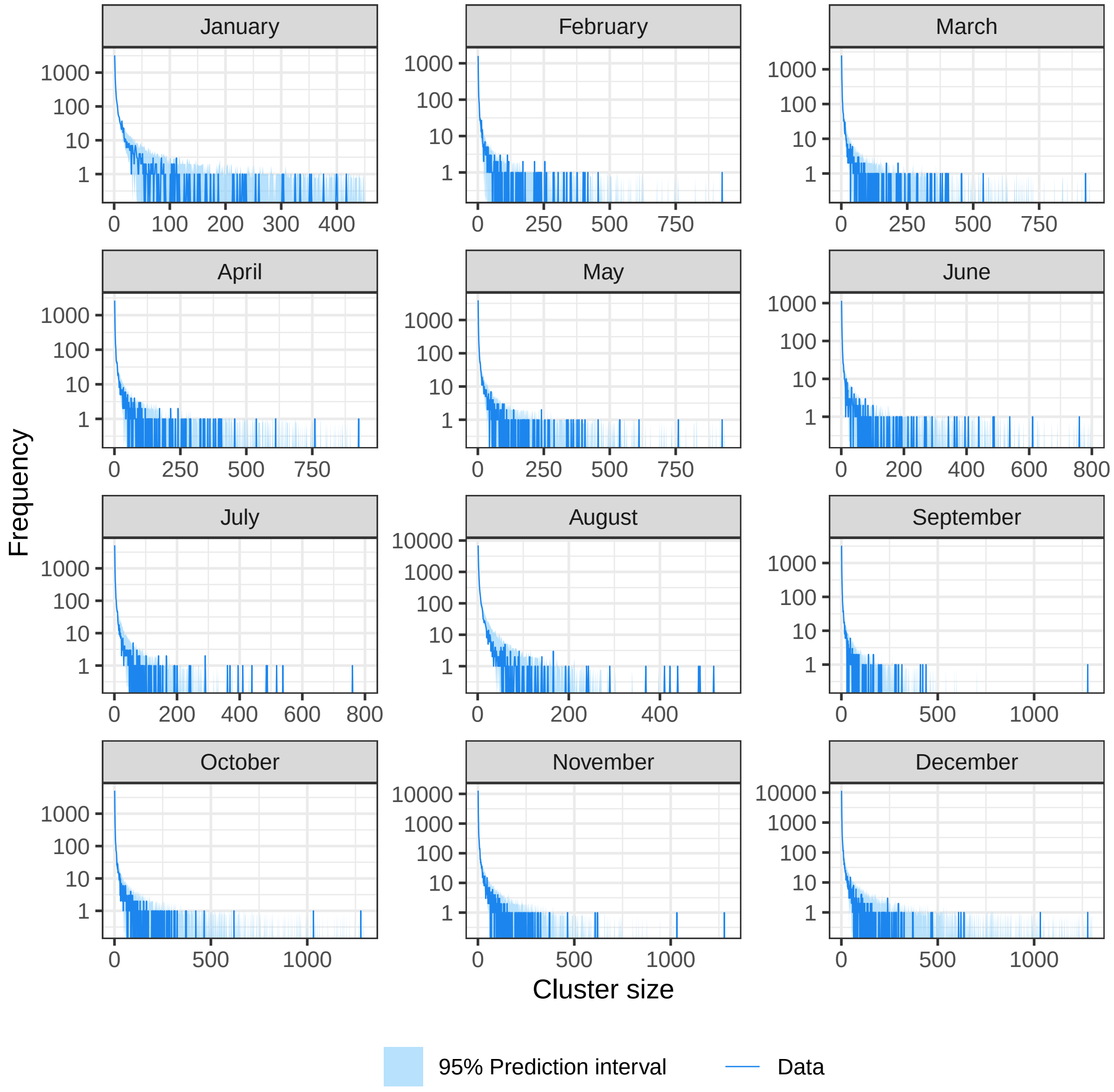

**Figure S12. Posterior predictive check of simulation of cluster size distribution (Denmark).**

#### 9.3 Germany

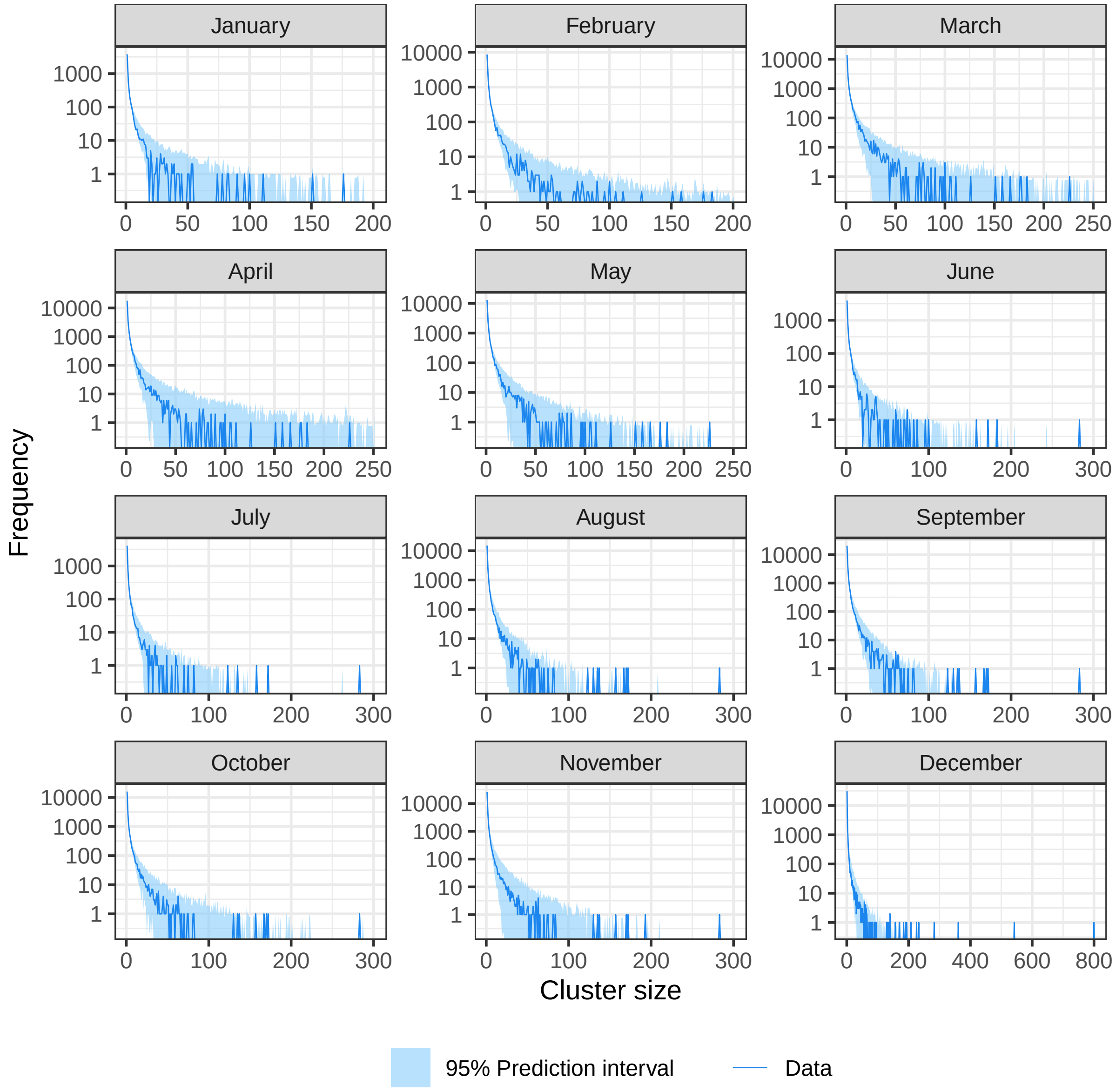

**Figure S13. Posterior predictive check of simulation of cluster size distribution (Germany).**

### 10 Goodness of fit

We performed a goodness of fit check to assess whether our main model of the cluster size distribution (presented in the paper) is compatible with the cluster data from Switzerland, Denmark and Germany. For each country and month we took the estimated mean of the effective reproduction number $R_e$, the dispersion parameter $k$, the mutation probability $\mu$ and the testing probability $\tau_{test}$ as an input for our model of the cluster size distribution. This line is surrounded by an area whose boundaries are the results of the model of the cluster size distribution when the estimated 2.5% and 97.5% quantiles of the estimated posterior distributions of the parameters are used as input. The following figures present the outcome of the goodness of fit check.

#### 10.1 Switzerland

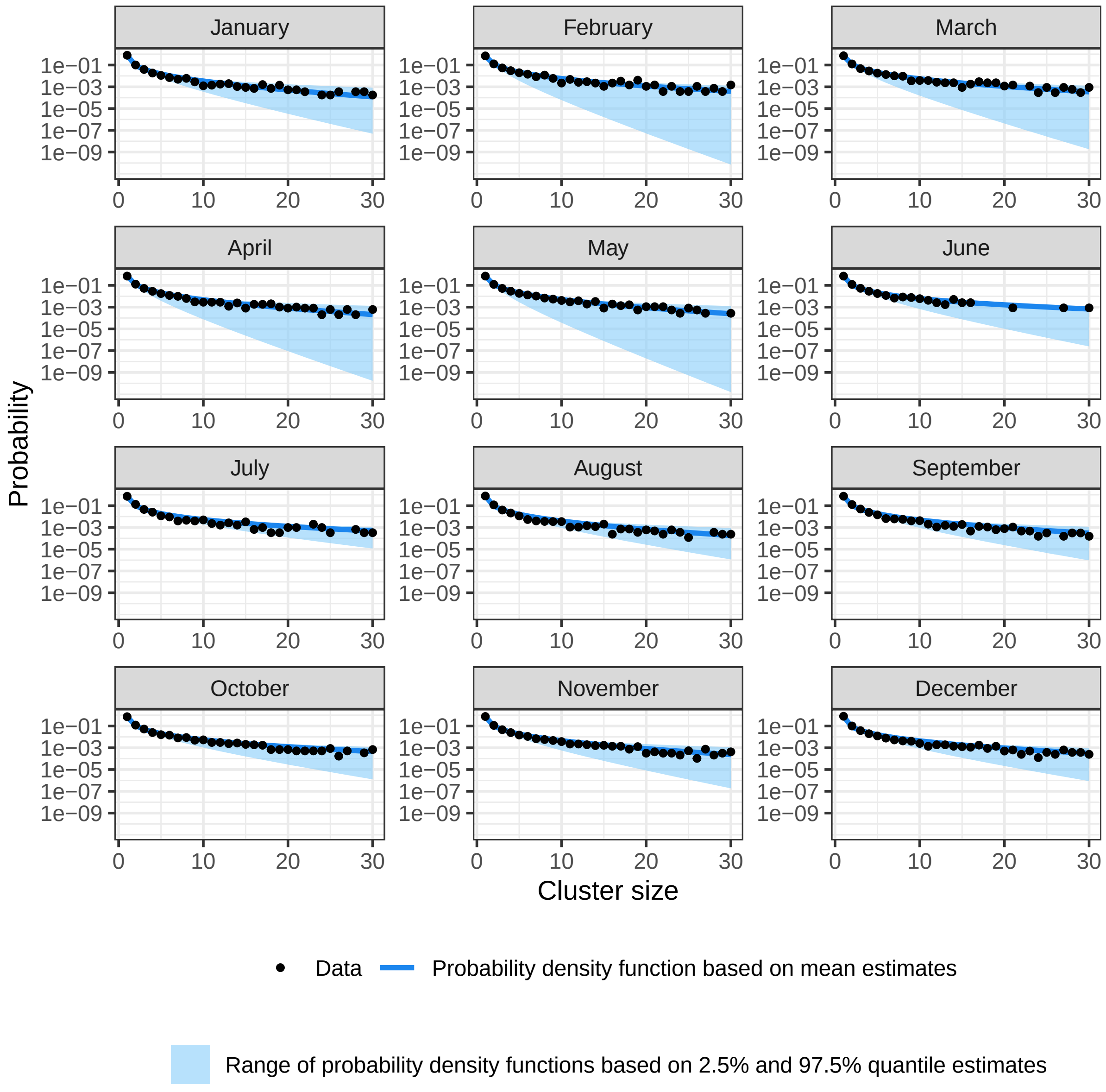

**Figure S14. Goodness of fit of cluster size distribution (Switzerland).**

#### 10.2 Denmark

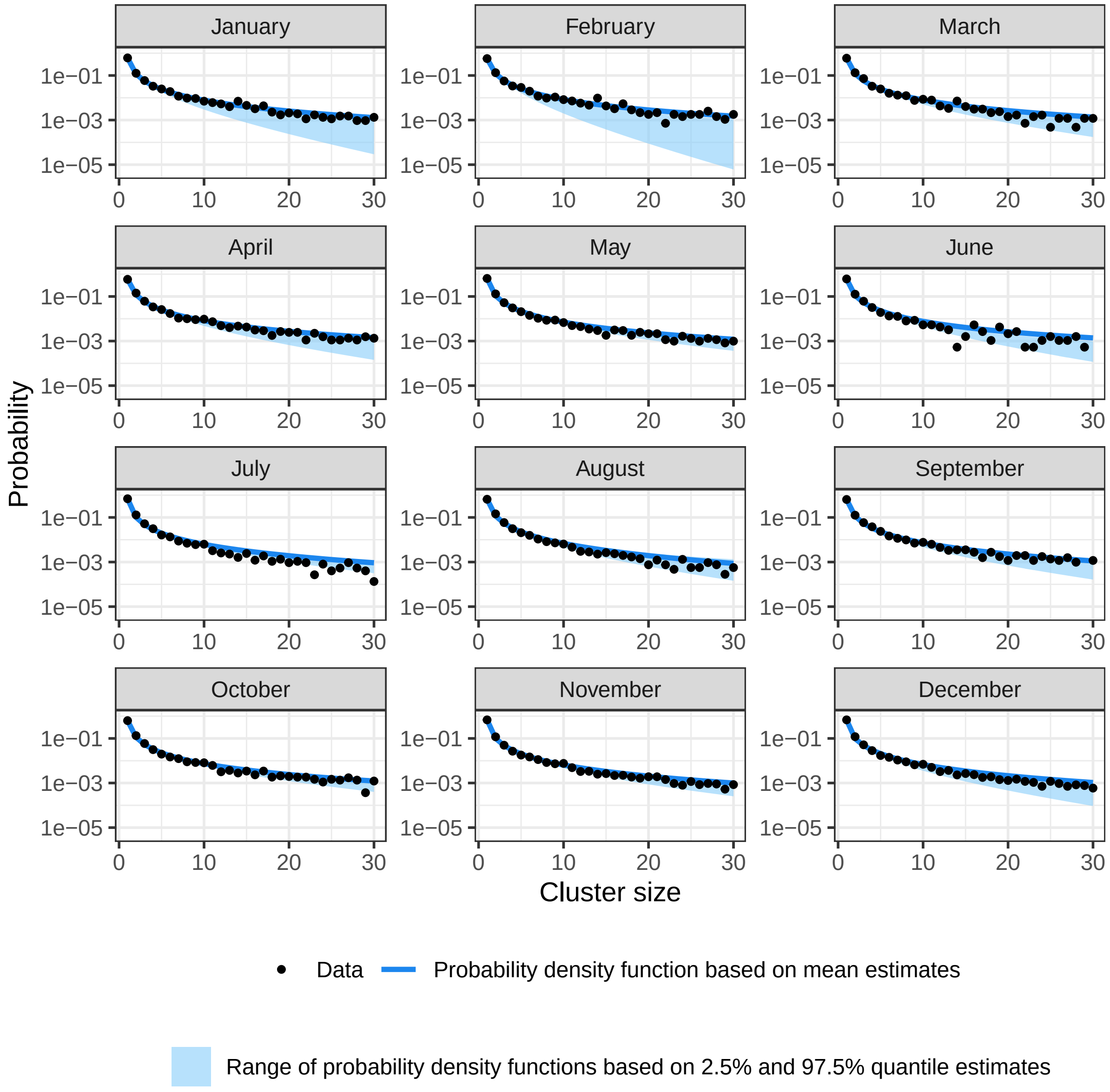

**Figure S15. Goodness of fit of cluster size distribution (Denmark).**

#### 10.3 Germany

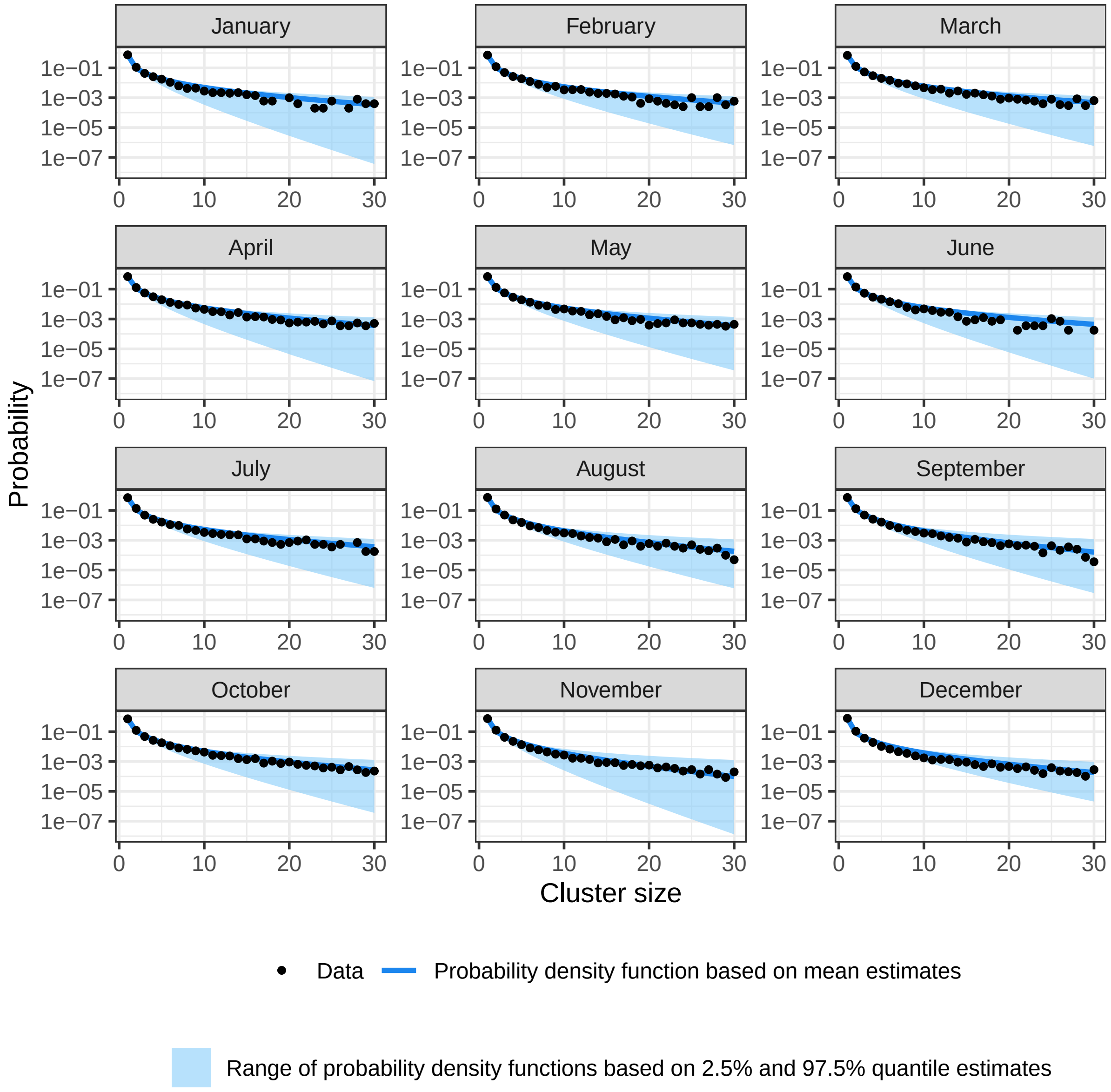

**Figure S16. Goodness of fit of cluster size distribution (Germany).**
