## Supplementary material for "Estimating *R_e_* and overdispersion in secondary cases from the size of identical sequence clusters of SARS-CoV-2": GISAID sequences for Switzerland

### SUPPLEMENTAL TABLE

**Data Availability**

GISAID Identifier: EPI_SET_240326pm
doi: 10.55876/gis8.240326pm

All genome sequences and associated metadata in this dataset are published in GISAID's EpiCoV database. To view the contributors of each individual sequence with details such as accession number, Virus name, Collection date, Originating Lab and Submitting Lab and the list of Authors, visit 10.55876/gis8.240326pm

**Data Snapshot**

- EPI_SET_240326pm is composed of 96,611 individual genome sequences.
- The collection dates range from 2020-08-14 to 2022-03-23;
- Data were collected in 2 countries and territories;
- All sequences in this dataset are compared relative to hCoV-19/Wuhan/WIV04/2019 (WIV04), the official reference sequence employed by GISAID (EPI_ISL_402124). Learn more at https://gisaid.org/WIV04.
